# Ranking the effectiveness of worldwide COVID-19 government interventions

**DOI:** 10.1101/2020.07.06.20147199

**Authors:** Nils Haug, Lukas Geyrhofer, Alessandro Londei, Elma Dervic, Amélie Desvars-Larrive, Vittorio Loreto, Beate Pinior, Stefan Thurner, Peter Klimek

**Author notes:** Equal contributions (Nils Haug, Lukas Geyrhofer, Alessandro Londei).

## Abstract

Assessing the effectiveness of Non-Pharmaceutical Interventions (NPIs) to mitigate the spread of SARS-CoV-2 is critical to inform future preparedness response plans. Here we quantify the impact of 6,068 hierarchically coded NPIs implemented in 79 territories on the effective reproduction number, *R*_*t*_, of COVID-19. We propose a novel modelling approach that combines four computational techniques merging for the first time statistical, inference and artificial intelligence tools. We validate our findings with two external datasets with 48,000 additional NPIs from 226 countries. Our results indicate that a suitable combination of NPIs is necessary to curb the spread of the virus. Less intrusive and costly NPIs can be as effective as more intrusive, drastic, ones, e.g., a national lockdown. Using country-specific what-if scenarios we assess how the effectiveness of NPIs depends on the local context such as timing of their adoption, opening the way for forecasting the effectiveness of future interventions.

## 1 Introduction

In the absence of vaccines and antiviral medication, non-pharmaceutical interventions (NPIs) implemented in response to epidemic respiratory viruses are the only option to delay and moderate the spread of the virus in a population ^1^.

Confronted with the worldwide COVID-19 epidemic, most governments have implemented bundles of highly restrictive, sometimes intrusive NPIs. Decisions had to be taken under rapidly changing epidemiological situations, despite (at least in the very beginning of the epidemic) a lack of scientific evidence on the individual and combined effectiveness of these measures ^2–4^, degree of compliance of the population, and societal impact.

Government interventions may cause substantial economic and social costs ^5^ as well as affect individuals’ behaviour, mental health and social security ^6^. Therefore, knowledge on the most effective NPIs would allow stakeholders to judiciously and timely implement a specific sequence of key interventions to combat a potential resurgence of COVID-19 or any other future respiratory outbreak. As many countries rolled out several NPIs simultaneously, the challenge of disentangling the impact of each individual intervention arises.

To date, studies of the country-specific progression of the COVID-19 pandemic ^7^ have mostly explored the independent effects of a single category of interventions. These categories include travel restrictions ^2, 8^, social distancing ^9–12^, or personal protective measures ^13^. Some studies focused on a single country or even a town ^14–18^. Other research combined data from multiple countries but pooled NPIs into rather broad categories ^15, 19–21^, which eventually limits the assessment of specific, potentially critical, NPIs, that may be less costly and more effective than others. Despite their widespread use, relative ease of implementation, broad choice of available tools, and their importance in developing countries where other measures (e.g., increases in healthcare capacity, social distancing, or enhanced testing) are difficult to implement ^22^, little is currently known about the effectiveness of different risk communication strategies. One reason for this knowledge gap might be that many NPI trackers do not clearly code only-communication actions or cover such measures rather superficially. For example, the WHO dataset ^23^ and the CoronaNet dataset ^24^ both report communication strategies (or public awareness measures) in two broad categories. However, an accurate assessment of communication activities requires information on the targeted public, means of communication and content of the message. Other government communications are sometimes summarized in non-communication categories (e.g., communication on social distancing are included in “Social distancing” measures in the CoronaNet dataset and an extra data element specifies the degree of compliance). Additionally, modelling studies typically focus on NPIs that directly influence contact probabilities (e.g., social distancing ^12, 18^, self-isolation ^20^, etc.).

Using a comprehensive, hierarchically coded, data set of 6,068 NPIs implemented in 79 territories ^25^, here we analyse the impact of government interventions on *R*_*t*_, using harmonised results from a new multi-method approach consisting of (i) a case-control analysis (CC), (ii) a step function approach to LASSO time-series regression (LASSO), (iii) random forests (RF) and (iv) Transformers (TF). We contend that the combination of four different methods, combining statistical, inference and artificial intelligence classes of tools, allows to also assess the structural uncertainty of individual methods ^26^. We also investigate country-specific control strategies as well as the impact of some selected country-specific metrics.

All approaches (i-iv) yield comparable rankings of the effectiveness of different categories of NPIs across their hierarchical levels. This remarkable agreement allows us to identify a consensus set of NPIs that lead to a significant reduction of *R*_*t*_. We validate this consensus set using two external datasets covering 42,151 measures in 226 countries. Further, we evaluate the heterogeneity of the effectiveness of individual NPIs in different territories. We find that time of implementation, already implemented measures, different governance indicators ^27^, as well as human and social development affect the effectiveness of NPIs in the countries to varying degrees.

## 2 Results

### Global approach

Our main results are based on the CSH COVID-19 Control Strategies List (CCCSL) ^25^. This data set provides a hierarchical taxonomy of 6,068 NPIs, coded on four levels, including eight broad themes (level 1, L1) are divided into 63 categories of individual NPIs (level 2, L2) that include >500 subcategories (level 3, L3) and >2,000 codes (level 4, L4). We first compare the results for the NPIs’ effectiveness rankings for the four methods of our approach (i-iv) on L1 (themes); see SI Figure S1. A clear picture emerges where the themes of social distancing and travel restrictions are top-ranked in all methods, whereas environmental measures (e.g., cleaning and disinfecting shared surfaces) are ranked least effective.

We next compare results obtained on L2 of the NPI data set, i.e., using the 46 NPI categories implemented more than five times. The methods largely agree on the list of interventions that have a significant effect on *R*_*t*_, see Figure 1 and Table 1. The individual rankings are highly correlated with each other (*p* = 0.0008, see Methods). Six NPI categories show significant impacts on *R*_*t*_ in all four methods. In Table S3 we list the subcategories (L3) belonging to these consensus categories.

**Table 1:** Comparison of effectiveness rankings on L2. Out of the 46 different NPI categories, all four methods show significant results for six NPIs (consensus 4); three methods agree on 14 further NPIs (consensus 3). We report the average normalised score, the observed reduction in *R*_*t*_ for the various methods and the NPI importance for the random forest. The numbers in brackets give half of the amount by which the last digit of the corresponding number outside the brackets fluctuates within the 95% confidence interval.

| L2 category | Score | Consensus | $\Delta R_t^{CC}$ | $\Delta R_t^{LASSO}$ | Importance (RF) | $\Delta R_t^{TF}$ |
| --- | --- | --- | --- | --- | --- | --- |
| Small gathering cancellation | 83% | 4 | -0.35 (2) | -0.22 (5) | 0.020 (2) | -0.327 (3) |
| Closure of educational institutions | 73% | 4 | -0.16 (2) | -0.21 (4) | 0.028 (2) | -0.146 (2) |
| Border restriction | 56% | 4 | -0.23 (2) | -0.12 (2) | 0.017 (2) | -0.057 (2) |
| Increase availability of personal protective equipment (PPE) | 51% | 4 | -0.11 (2) | -0.13 (2) | 0.012 (1) | -0.062 (2) |
| Individual movement restrictions | 42% | 4 | -0.13 (2) | -0.08 (3) | 0.017 (2) | -0.121 (2) |
| National lockdown | 25% | 4 | -0.14 (3) | -0.09 (2) | 0.0020 (9) | -0.008 (3) |
| Mass gathering cancellation | 53% | 3 | -0.33 (2) | 0 | 0.012 (1) | -0.127 (2) |
| Educate and actively communicate with the public | 48% | 3 | -0.18 (4) | 0 | 0.018 (2) | -0.276 (2) |
| The government provides assistance to vulnerable populations | 41% | 3 | -0.17 (3) | -0.18 (4) | 0.009 (1) | 0.090 (3) |
| Actively communicate with managers | 40% | 3 | -0.15 (2) | -0.20 (4) | 0.004 (2) | -0.050 (2) |
| Measures for special populations | 37% | 3 | -0.19 (2) | 0 | 0.008 (1) | -0.100 (2) |
| Increase healthcare workforce | 35% | 3 | -0.17 (20) | -0.13 (3) | 0.030 (8) | 0.011 (2) |
| Quarantine | 30% | 3 | -0.28 (2) | -0.2 (1) | 0.0023 (9) | 0.023 (2) |
| Activate or establish emergency response | 29% | 3 | -0.13 (2) | 0 | 0.0037 (9) | -0.121 (2) |
| Enhance detection system | 25% | 3 | -0.19 (3) | 0 | 0.0032 (9) | -0.106 (2) |
| Increase in medical supplies and equipment | 25% | 3 | -0.13 (3) | -0.004 (3) | 0.003 (2) | -0.200 (3) |
| Police and army interventions | 23% | 3 | -0.16 (2) | 0 | 0.003 (2) | -0.091 (2) |
| Travel alert and warning | 20% | 3 | -0.13 (3) | 0.0 (1) | 0.002 (1) | -0.159 (3) |
| Public transport restriction | 13% | 3 | -0.20 (4) | -0.01 (7) | 0.004 (1) | -0.023 (3) |
| Actively communicate with healthcare professionals | 11% | 3 | 0 | -0.08 (4) | 0.003 (1) | -0.003 (2) |

**Figure 1:**
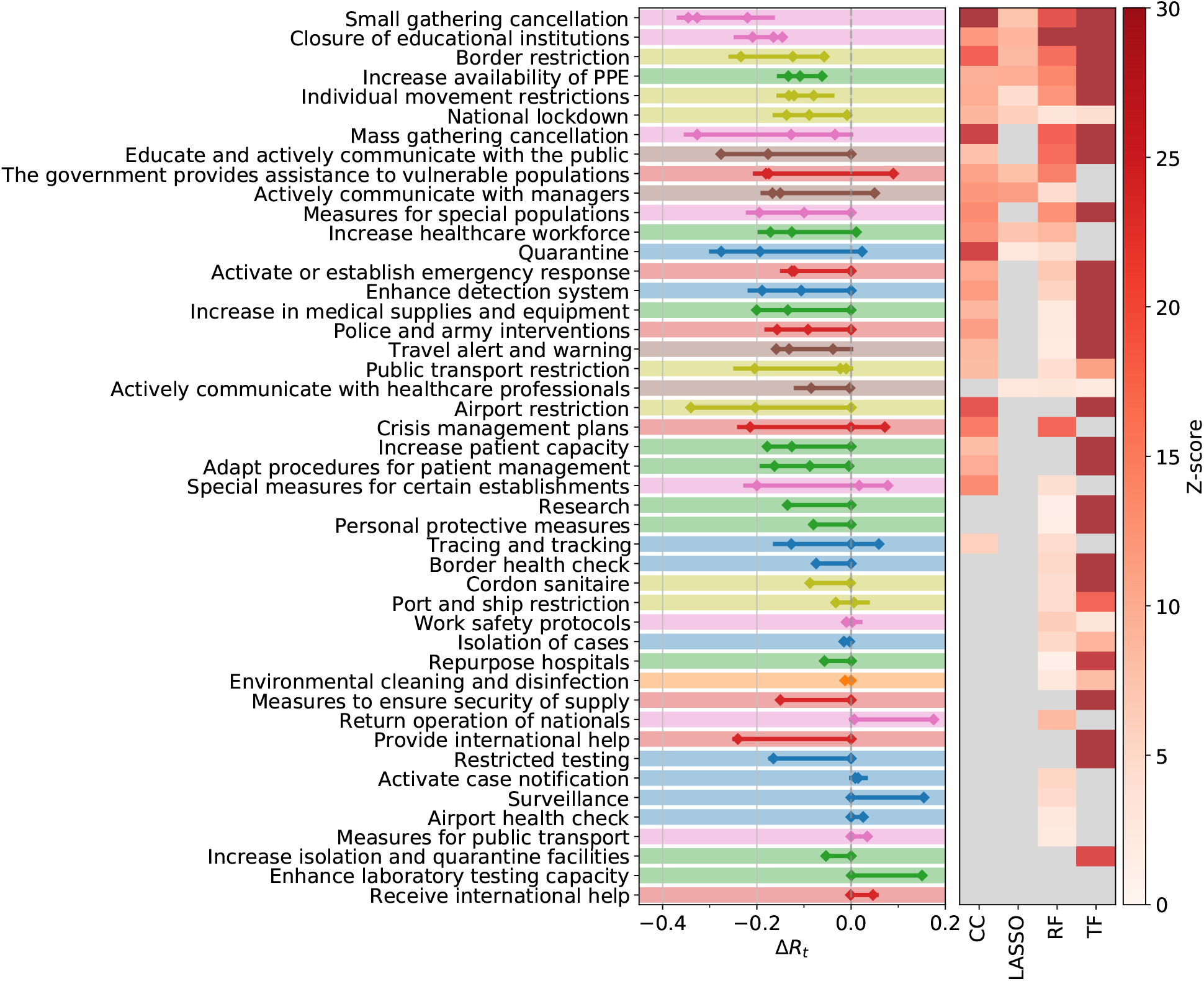
Decrease in the effective reproduction number, Δ*R*_*t*_, for 46 NPIs at L2, as quantified by case-control analysis (CC), LASSO, and the transformer (TF) regression. The left panel shows the combined 95% confidence interval of Δ*R*_*t*_ for the most effective interventions across all included territories. The heatmap in the right panel shows the corresponding *Z*-scores of the measure effectiveness as determined by the four different methods. Gray color indicates no significantly positive effect. NPIs are ranked according to the number of methods agreeing on their impacts, from top (significant in all methods) to bottom (ineffective in all analyses). L1 themes are colour-coded as in Figure S1.

A normalised score for each NPI category is obtained by rescaling the result of each method to range between zero (least effective) and one (most effective) and then averaging this score. The maximal (minimal) NPI score is therefore 100% (0%), meaning that the measure is the most (least) effective measure in each method. Amongst the six full consensus NPI categories, the largest impacts on *R*_*t*_ are displayed by small gathering cancellations (83%, Δ*R*_*t*_ between −0.22 and −0.35), the closure of educational institutions (with a score of 73% and estimates for Δ*R*_*t*_ ranging from −0.15 to −0.21), and border restrictions (56%, Δ*R*_*t*_ between −0.057 and −0.23). The consensus measures also include NPIs aiming to increase healthcare and public health capacities (increase availability of personal protective equipment (PPE): 51%, Δ*R*_*t*_ −0.062 to −0.13), individual movement restrictions (42%, Δ*R*_*t*_ −0.08 to −0.13) and national lockdown (including stay-at-home order in US states) (25%, Δ*R*_*t*_ −0.008 to −0.14).

We find fourteen additional NPI categories consensually in three of our methods. These include mass gathering cancellations (53%, Δ*R*_*t*_ between −0.13 and −0.33), risk communication activities to inform and educate the public (48%, Δ*R*_*t*_ between −0.18 and −0.28), and government assistance to vulnerable populations (41%, Δ*R*_*t*_ between −0.17 and −0.18).

Amongst the least effective interventions we find: government actions to provide or receive international help, measures to enhance testing capacity or improve case detection strategy (which can be expected to lead to a short-term rise in cases), tracing and tracking measures, as well as land border and airport health checks and environmental cleaning.

In Figure 2 we visualise the findings on the NPIs’ effectiveness in a co-implementation network. Nodes correspond to categories (L2) with a size being proportional to their normalised score. Directed links from *i* to *j* indicate a tendency that countries implement NPI *j* after they implemented *i*. The network therefore illustrates the typical NPI implementation sequence in the 56 countries and the steps within this sequence that contribute most to a reduction of *R*_*t*_. For instance, there is a pattern where countries first cancel mass gatherings before moving on to cancellations of specific types of small gatherings, where the latter associates on average with more substantial reductions in *R*_*t*_. Education and active communication is one of the most effective “early measures” (implemented around 15 days before 30 cases were reported and well before the majority of other measures comes). Most social distancing (i.e., closure of educational institutions), travel restriction measures (i.e., individual movement restrictions like curfew, national lockdown) and measures to increase the availability of PPE are typically implemented within the first two weeks after reaching 30 cases with varying impacts on the *R*_*t*_; see also Figure 1.

**Figure 2:**
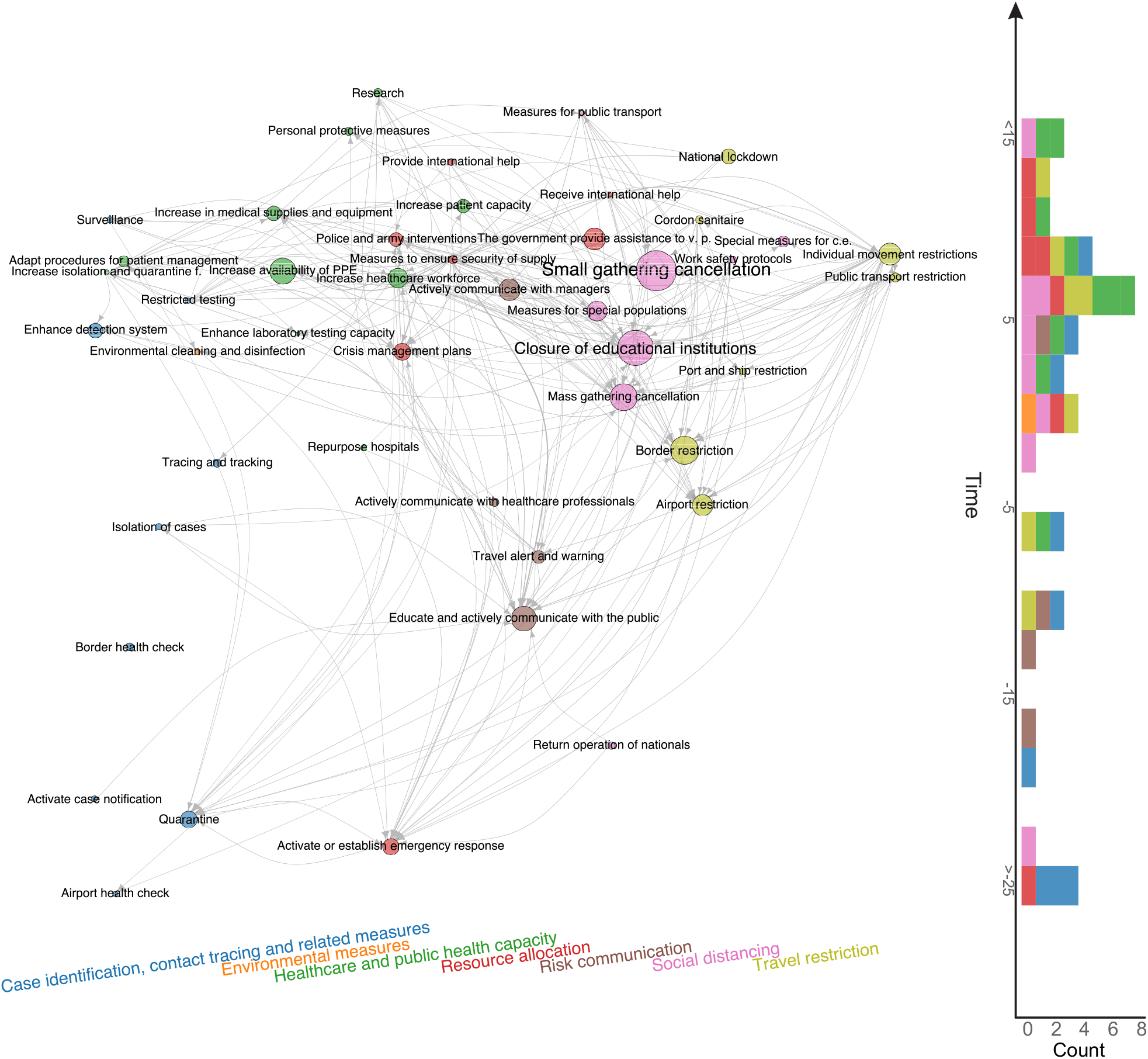
Time-ordered NPI co-implementation network across countries. Nodes are categories (L2) with colour indicating the theme (L1) and size being proportional to the average effectiveness of the intervention. Arrows from nodes *i* to *j* represent that countries which have already implemented intervention *i* tend to implement intervention *j* later in time. Nodes are positioned vertically according to their average time of implementation (measured relative to the day where the country reached 30 confirmed cases) and horizontally according to their L1 theme.

Within the CC approach, we can further explore these results on a finer hierarchical level. We show results for 18 NPIs (L3) of the risk communication theme in the SI; see Table S2. The most effective communication strategies include warnings against travel to and return from high risk areas (Δ*R*_*t*_ = −0.14(1)) and several measures to actively communicate with the public.

These include to encourage, e.g., staying at home (Δ*R*_*t*_ = −0.14(1)), social distancing (Δ*R*_*t*_ = −0.20(1)), workplace safety measures (Δ*R*_*t*_ = −0.18(2)), self-initiated isolation of people with mild respiratory symptoms (Δ*R*_*t*_ = −0.19(2)) as well as information campaigns (Δ*R*_*t*_ = −0.13(1)) (through various channels such as press, flyers, social media, or phone messages).

### Validation with external datasets

We validate our findings with results from two external datasets, see Methods. In the WHO-PHSM dataset ^23^ we find seven full-consensus measures (agreement on significance by all methods) and 17 further measures with three agreements, see SI Figure S27. These consensus measures show a large overlap with the consensus measures (three or four matches in our methods) identified using the CCCSL and includes as top-ranked NPI measures aiming at strengthening the healthcare system and the testing capacity (labeled as “Scaling up”), e.g., increase healthcare workforce, purchase of medical equipment, tests, masks, financial support to hospitals, increase patient capacity, increase domestic production of PPE). Other consensus measures consist of social distancing measures (“Cancelling, restricting or adapting private gatherings outside the home”, adapting or closing “offices, businesses, institutions and operations”, “cancelling, restricting or adapting mass gatherings”), measures for special populations (“protecting population in closed settings”, encompassing long-term care facilities and prisons), school closures, (international and domestic) travel restrictions (stay-at-home order – equivalent to confinement in the WHO-PHSM coding – restricting entry and exit, travel advice and warning, “closing international land borders”, “entry screening and isolation or quarantine)”. “Wearing a mask” exhibits a significant impact on *R*_*t*_ in three methods (Δ*R*_*t*_ between −0.018 and −0.12). The consensus measures also include financial packages and general public awareness campaigns (as part of “Communications and engagement” actions). The least effective measures include active case detection, contact tracing, as well as environmental cleaning and disinfection.

The CCCSL results are also compatible with findings from the CoronaNet dataset ^24^; see SI Figures S28–S29. Analyses show four full-consensus measures and 13 further NPIs with an agreement of three methods. These consensus measures include general social distancing measures (no specific coding available), restriction and regulation of non-essential businesses, restrictions of mass gatherings, closure and regulation of schools, travel restrictions (e.g., internal and external border restrictions, curfew), measures aiming to increase healthcare workforce (e.g., “Nurses”, “Unspecified health staff”) and medical equipment (e.g., PPE, “Ventilators”, “Unspecified health materials”), quarantine (i.e., voluntary or mandatory self-quarantine and quarantine at a government hotel or facility), and measures to increase public awareness (“Disseminating information related to COVID-19 to the public that is reliable and factually accurate”).

Twenty-three NPIs in the CoronaNet dataset do not show statistical significance in any method, including several restrictions and regulations of government services (e.g., for tourist sites, parks, public museums, telecommunications), hygiene measures for public areas, and other measures that target very specific populations (e.g., certain age groups, visa extensions).

### Country-level approach

A sensitivity check of our results with respect to the removal of individual continents from the analysis also indicates substantial variations between world geographical regions in terms of NPI effectiveness (see SI). To further quantify how much the effectiveness of an NPI depends on the particular territory (country or US state) where it has been introduced, we measure the heterogeneity of the NPI rankings in different territories through an entropic approach in the transformer (TF) method; see Methods. Figure 3 shows the normalised entropy of each NPI category versus its rank. A value of entropy close to zero implies that the corresponding NPI has a similar rank relative to all other NPIs in all territories. In other words, the effectiveness of the NPI does not depend on the specific country or state. On the other hand, a high value of the normalised entropy signals that the performance of each NPI depends largely on the geographical region.

**Figure 3:**
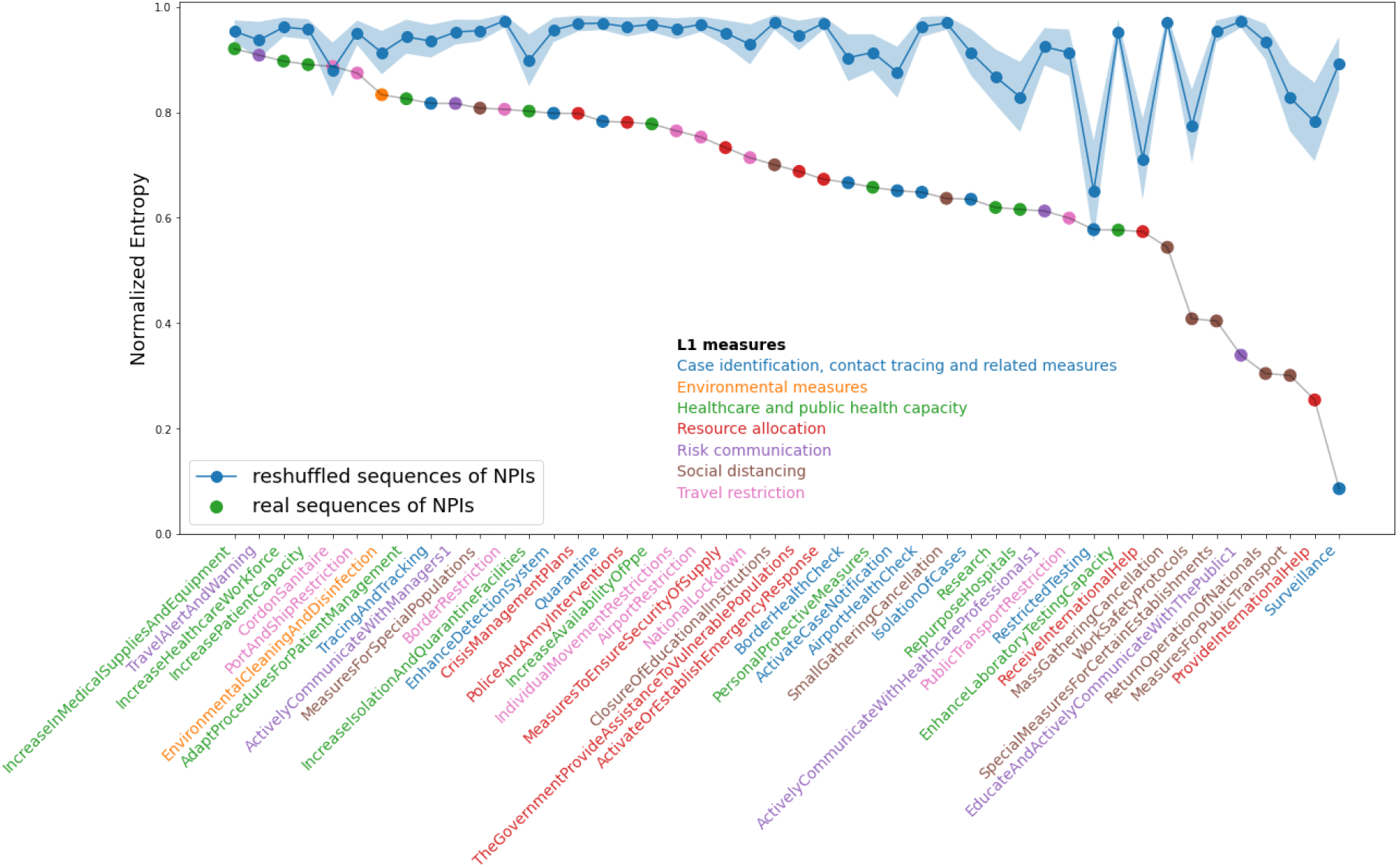
Normalised entropies vs rank for all the NPIs at level L2. Each NPI name is coloured according to its theme of belonging (L1) as indicated in the legend. The blue curve represents the same information obtained out of a reshuffled data set of NPIs.

The values of the normalised entropies for many NPIs are far from being one and below the corresponding values obtained through a temporal reshuffling of the NPIs in each country. The effectiveness of many NPIs therefore is, first, significant and, second, heavily dependent on the local context, which is a combination of socio-economic features and NPIs already adopted. In general, social distancing measures and travel restrictions show a high entropy (effectiveness varies a lot across countries) whereas case identification, contact tracing and healthcare measures show substantially less country dependence.

We further explore this interplay of NPIs with socio-economic factors by analysing the effects of demographic and socio-economic covariates, as well as indicators for governance, human and economic development in the CC method (see SI). While the effects of most indicators vary across different NPIs at rather moderate levels, we find a robust tendency that NPI effectiveness correlates negatively with indicator values for governance-related accountability and political stability (as quantified by World Governance Indicators provided by the World Bank).

The heterogeneity of the effectiveness of individual NPIs across countries points to a non-independence among the different NPIs, therefore the impact of a specific NPI cannot be evaluated in isolation. Instead, one has to look at the combination of NPIs adopted in a particular country. Since it is not possible in the real world to change the sequence of NPIs adopted, we resort to what-if experiments to identify the most likely outcome of an artificial sequence of NPIs in each specific country. Within the TF approach, we selectively knock-out one NPI at the time from all the sequences of interventions in all countries and compute the ensuing evolution of *R*_*t*_ compared to the actual case.

To quantify whether the effectiveness of a specific NPI depends on its epidemic age of implementation, we study artificial sequences of NPIs constructed by shifting the selected NPI to other days, keeping the other NPIs fixed. In this way, for each country and each NPI, we obtain a curve of the most likely change of *R*_*t*_ vs the adoption time of the specific NPI.

Figure 4 reports an example of the results for a selection of NPIs (we refer to the SI for a larger report about other NPIs). Each curve shows the average change of *R*_*t*_ vs the adoption time of the NPI, averaged over the countries where the NPI has been adopted. Panel A refers to the national lockdown (including stay-at-home order implemented in the US states). Our results show a moderate effect of this NPI (low change in *R*_*t*_) as compared to other, less drastic measures. Panel B shows NPIs with a “the earlier, the better” pattern. For those measures (“Closure of educational institutions”, “Small gatherings cancellation”, “Airport restrictions” and many more in the SI) early adoption is always more beneficial. Panel C, “Enhancing testing capacity” and “Surveillance”, exhibit a negative impact (i.e., an increase) on *R*_*t*_ presumably related to the fact that more testing allows for surfacing more cases. Finally, Panel D, showing “Tracing and tracking” and “Activate case notification”, display an initially negative effect that turns positive (i.e., toward a reduction of *R*_*t*_). We refer to the Supplementary Information for a more comprehensive analysis of all the NPIs.

**Figure 4:**
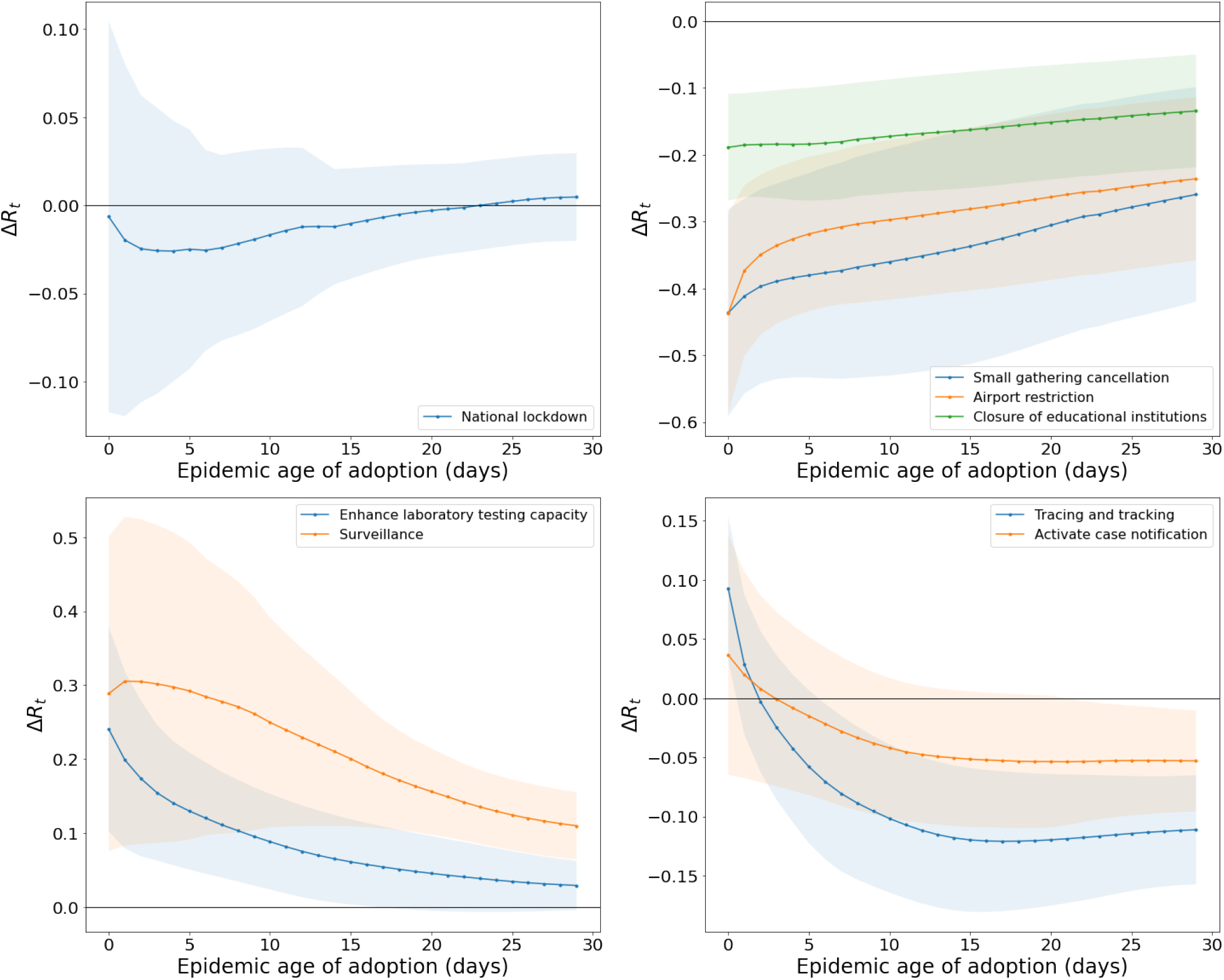
Change of *R*_*t*_ as a function of the adoption time of the NPI, averaged over the countries where the NPI has been adopted. Negative (Positive) values here mean that the adoption of the NPI has reduced (increased) the value of *R*_*t*_. Panel A: “National lockdown” (including “stay-at-home Order in the US states). Panel B: A selection of NPIs that display the “The earlier the better” behaviour, i.e., their impact is better if implemented at earlier epidemic ages. Panel C: “Enhancing testing capacity” and “Surveillance”. Panel D: “Tracing and Tracking” and “Activate case notification”.

## 3 Discussion

Our study dissects the entangled packages of NPIs ^25^ and quantifies their effectiveness. We validate our findings using three different datasets and four independent methods. Our findings suggest that no NPI acts as a silver bullet on the spread of COVID-19. Instead, we identify several decisive interventions that significantly contribute to reducing *R*_*t*_ below one and should therefore be considered to efficiently flatten the curve facing a potential second COVID-19 wave or any similar future viral respiratory epidemics.

The most effective NPIs include curfews, lockdowns, and closing and restricting places where people gather in smaller or large numbers for an extended period of time. This includes small gathering cancellations (closures of shops, restaurants, gatherings of 50 persons or less, mandatory home office, etc.) and closure of educational institutions. While in previous studies based on smaller numbers of countries, school closures had been attributed a little effect on the spread of COVID-19 ^19, 20^, more recent evidence has been in favour of the importance of this NPI ^28, 29^. School closures in the US have been found to reduce COVID-19 incidence and mortality by about 60% ^28^. This result is also in line with a contact tracing study from South Korea, which identified adolescents aged 10–19 as more likely to spread the virus than adults and children in household settings ^30^. Individual movement restrictions (including curfew, the prohibition of gatherings and movements for non-essential activities or measures segmenting the population) were also amongst the top-ranked measures.

However, such radical measures present with adverse consequences. School closure interrupts learning, can lead to poor nutrition, stress and social isolation in children ^31–33^. Home confinement has strongly increased the rate of domestic violence in many countries, with a huge impact on women and children ^34, 35^, while it has also limited the access to long-term care, such as chemotherapy, with significant impacts on patients’ health and survival chance ^36, 37^. Governments may have to look towards less stringent measures, encompassing a maximum of effective prevention but enabling an acceptable balance between benefits and drawbacks ^38^.

Previous statistical studies on the effectiveness of lockdowns came to mixed conclusions. Whereas a relative reduction of *R*_*t*_ of 5% was estimated using a Bayesian hierarchical model ^19^, a Bayesian mechanistic model estimated a reduction of 80% ^20^, though some questions have been raised regarding the latter work ^26^. Our results point to a mild impact of them due to an overlap with effects of other measures adopted earlier and included in what is referred to as “national (or full) lockdown”. Indeed, the national lockdown encompasses multiple NPIs (e.g., closure of land, sea and air borders, schools, non-essential shops, prohibition of gatherings, of visiting nursing homes) that countries may have already adopted. From this perspective, the relatively attenuated impact of the national lockdown is explained as the little delta after other concurrent NPIs have been adopted. This conclusion does not rule out the effectiveness of an early national lockdown but suggests that a suitable combination (sequence and time of implementation) of a smaller package of such measures *can* substitute a full lockdown in terms of effectiveness while reducing adverse impacts on the society, economy, humanitarian response system, and the environment ^6, 39–41^.

Taken together, the social distancing and movement restriction measures discussed above can therefore be seen as the “nuclear option” of NPIs: highly effective but causing substantial collateral damages on society, the economy, trade, and human rights ^4, 39^.

We find strong support for the effectiveness of border restrictions. The role of travelling in the global spread of respiratory diseases has proved central during the first SARS epidemic (2002-2003) ^42^, but travelling restrictions show a large impact on trade, economy, and humanitarian response system globally ^41, 43^. The effectiveness of social distancing and travel restrictions is also in line with results from other studies, which used different statistical approaches, epidemiological metrics, geographic coverage, and classifications of NPIs ^2, 8–11, 13, 19, 20^.

We also find a number of highly effective NPIs that can be considered to be less costly. For instance, we find that risk communication strategies feature prominently amongst consensus NPIs. This includes government actions intended to educate and actively communicate with the public. To the best of our knowledge, our study provides the first quantitative evidence for the effectiveness of such measures. The effective policies include encouraging staying at home, promoting social distancing and workplace safety measures, encouraging the self-initiated isolation of people with symptoms, travel warnings, as well as information campaigns (mostly via social media). All these measures are non-binding government advice, contrasting with the mandatory border restriction and social distancing measures that are often enforced by police or army interventions and sanctions. Surprisingly, communicating on the importance of social distancing has been only marginally less effective than imposing distancing measures by law. The publication of guidelines and work safety protocols to managers and healthcare professionals was also associated with a reduction of *R*_*t*_, suggesting that communication efforts also need to be tailored toward key stakeholders. Communication strategies aim at empowering communities with correct information about COVID-19. Such measures can be of crucial importance to target specific demographic strata found to play a dominant role in driving the COVID-19 spread (e.g., communication strategies to target individuals aged <40y ^44^)

Government food assistance programs and other financial supports for vulnerable populations (via taxation) also turned out to be highly effective. Such measures are, therefore, not only impacting the socio-economic sphere ^45^ but have also a positive effect on public health. For instance, facilitating people’s access to tests or allowing them to self-isolate without fear of losing their job, may help reducing the *R*_*t*_.

Some measures are ineffective in (almost) all methods and datasets, e.g., environmental measures to disinfect and clean surfaces and objects in public and semi-public places. This finding is at odds with current recommendations of the WHO for environmental cleaning in non-healthcare settings ^46^ and calls for a closer examination of the effectiveness of such measures. However, environmental measures (e.g., cleaning of shared surfaces, waste management, approval of a new disinfectant, increase ventilation) is seldom reported by governments or media, and therefore not collected by the NPI trackers, which could lead to an under-estimation of their impact. We also find no evidence for the effectiveness of social distancing measures in public transports. While infections on busses and trains have been reported ^47^, our results may suggest a limited contribution of such cases to the overall virus spread. A heightened public risk awareness associated with commuting (e.g., people being more likely to wear face masks) might contribute to this finding ^48^. However, we should notice that measures aiming a limiting engorgement or increasing distancing in public transports have been highly diverse (from complete cancellation of all public transports to increase in frequency of the traffic to reduce traveler density) and could therefore lead to largely different effectiveness, also depending on the local context.

The effectiveness of individual NPIs is heavily influenced by governance (see SI) and local context, as evidenced by the results of the entropic approach. This local context includes the stage of the epidemic, socio-economic, cultural and political characteristics, and other NPIs already implemented. By focusing on individual countries, the what-if experiments using artificial country-specific sequences of NPIs offer a novel way to quantify the importance of this local context with respect to measure effectiveness. Our main takeaway here is that one and the same NPI can have a drastically different impact if taken early or later, or in a different country.

It is interesting to comment on the impact that “Enhancing testing capacity” and “Tracing and tracking” would have had if adopted at different points in times. Counterintuitively, tracing, tracking and testing measures should display a short-term increase of *R*_*t*_ if they are effective, as more cases will be found. For countries implementing these measures early this is indeed what we find. However, countries implementing these NPIs later did not necessarily find more cases, as shown by the corresponding decrease in *R*_*t*_, We focused on March and April 2020, a period in which many countries had surged in positive cases that overwhelmed their testing and tracing capacities, which rendered the corresponding NPIs ineffective.

### Strengths & Limitations

The assessment of the effectiveness of NPIs is statistically challenging, as measures were typically implemented simultaneously and because their impact might well depend on the particular implementation sequence. Some NPIs appear in almost all countries whereas others only in few, meaning that we could miss some rare but effective measures due to a lack of statistical power. While some methods might be prone to overestimating effects from an NPI due to insufficient adjustments for confounding effects from other measures, other methods might underestimate the contribution of an NPI by assigning its impact to a highly correlated NPI. As a consequence, estimates of Δ*R*_*t*_ might vary substantially across different methods, whereas the agreement on the significance of individual NPIs is much more pronounced. The strength of our study, therefore, lies in the harmonization of these four independent methodological approaches, combined with the usage of an extensive data set on NPIs. This allows us to estimate the structural uncertainty of NPI effectiveness, i.e., the uncertainty introduced by choosing a certain model structure. Moreover, whereas previous studies often subsumed a wide range of social distancing and travel restriction measures under a single entity, our analysis contributes to a more fine-grained understanding of each NPI.

The CCCSL data set features non-homogeneous data completeness across the different territories and data collection could be biased by the data collector (native versus non-native) as well as the information communicated by governments. Moreover, the coding system presents some drawbacks, notably because some interventions could belong to more than one category but are only recorded once. Compliance with NPIs is crucial for their effectiveness, yet we assumed a comparable degree of compliance by each population. We tried to mitigate this issue by validating our findings on two external databases, even if those are subject to similar limitations. Additionally, we neither took into account the stringency of NPI implementation, and not all methods were able to describe potential variations of NPI effectiveness over time, besides the dependency on the epidemic age of its adoption.

To compute *R*_*t*_, we used time-series of the number of confirmed COVID-19 cases ^49^. This approach is likely to over-represent patients with severe symptoms and may be biased by variations in testing and reporting policies among countries. We assume a constant serial interval (average time-span between primary and secondary infection), however, this number shows considerable variations in the literature ^50^ and depends on measures such as social distancing and self-isolation.

## 4 Conclusions

Here we presented the outcome of an extensive analysis on the impact of 6,068 individual NPIs on the effective reproduction number *R*_*t*_ of COVID-19 in 79 territories worldwide. The adoption of the CCCSL data set ^25^ on NPIs and the use of two external validation datasets, encompassing together more than 48,000 NPIs over 226 countries, makes our study the largest on NPI effectiveness to date ^20, 21, 24, 51^.

The emerging picture reveals that no one-size-fits-all solution exists, and no single NPI can decrease *R*_*t*_ below one. Instead, in the absence of a vaccine or efficient anti-viral medication, a resurgence of COVID-19 cases can only be stopped by a suitable combination of NPIs, each tailored to the specific country and its epidemic age. These measures must be enacted in the optimal combination and sequence to be maximally effective on the spread of SARS-CoV-2 and thereby enable a faster re-opening.

We showed that the most effective measures include closing and restricting most places where people gather in smaller or larger numbers for extended periods of time (businesses, bars, schools, etc). However, we also find several highly effective measures that are less intrusive. These include land border restrictions, governmental support to vulnerable populations and risk communication strategies. We strongly recommend governments and other stakeholders to first consider the adoption of such NPIs, tailored to the local context, should infection numbers (re-)surge, before choosing the most intrusive options. Less drastic measures may also foster better compliance from the population.

Notably, the simultaneous consideration of many distinct NPI categories allows us to move beyond the simple evaluation of individual classes of NPIs to assess the collective impact of specific sequences of interventions instead. The ensemble of these results calls for a strong effort to simulate “what-if” scenarios at the country level for planning the most likely effectiveness of future NPIs, and, thanks to the possibility to go down to the level of individual countries and country specific circumstances, our approach is the first contribution to this end.

## Data Availability

The data set on non-pharmaceutical interventions to curb the spread of COVID-19 is available online via the following repository: https://github.com/amel-github/covid19-interventionmeasures. The data on confirmed cases of COVID-19 has been taken from https://github.com/CSSEGISandData/COVID-19.

## 5 Methods

### Data

#### NPI data

We use the publicly available Complexity Science Hub Vienna COVID-19 Control Strategies list (CCCSL) dataset on NPIs ^25^. Therein, NPIs are categorised using a four-level hierarchical coding scheme: L1 defines the theme of the NPI: “Case identification, contact tracing and related measures”, “Environmental measures”, “Healthcare and public health capacity”, “Resource allocation”, “Returning to normal life”, “Risk communication”, “Social distancing” and “Travel restriction”. Each L1 (theme) is composed of several categories (L2 of the coding scheme), that contain subcategories (L3) which are further subdivided to group codes (L4). The data set covers 56 countries; data for the USA is available at the state level (24 states). This makes a total of 79 territories. In this analysis, we use a static version of the CCCSL, retrieved on 17 August 2020, presenting 6,068 NPIs. A glossary of the codes is provided on github. For each country, we use the data until the day to which the measures have been reliably updated. NPIs that have been implemented in less than five territories are not considered, leading to a final number of 4,780 NPIs of 46 different L2 categories to be used in the analyses.

Secondly, we use the CoronaNet COVID-19 Government Response Event Dataset (v1.0) ^24^ that contains 31,532 interventions and covers 247 territories (countries and US states) (data extracted on 2020-08-17). For our analysis, we map their columns “type” and “type_sub_cat” onto L1 and L2, respectively. Definitions for the total 116 L2 categories can be found on the GitHub page of the project. Using the same criterion as for the CCCSL, we obtain a final number of 18,919 NPIs of 107 different categories.

Thirdly, we use the WHO Global Dataset of Public Health and Social Measures (thereafter called WHO-PHSM) ^23^ which merges and harmonizes the following data sets: ACAPS ^41^, Oxford COVID-19 Government Response Tracker ^52^, the Global Public Health Intelligence Network (GPHIN) of Public Health Agency of Canada (Ottawa, Canada), the CCCSL ^25^, the United States Centers for Disease Control and Prevention (CDC, Atlanta, USA) and the HIT-COVID data set ^53^. The WHO-PHSM Dataset contains 24,077 interventions and covers 264 territories (countries and US states) (data extracted on 2020-08-17). Their encoding scheme has a heterogeneous coding depth, and for our analysis we map “who_category” onto L1, and either take “who_subcategory” or a combination of “who_subcategory” and “who_measure” as L2. This results in 40 measure categories. A glossary is available at: https://www.who.int/emergencies/diseases/novel-coronavirus-2019/phsm.

#### COVID-19 case data

To estimate the effective reproduction number *R*_*t*_, and growth rates of the number of COVID-19 cases, we use time series of the number of confirmed COVID-19 cases in the 79 considered territories ^49^. To control for weekly fluctuations, we smooth the time series by computing the rolling average using a Gaussian window with a standard deviation of two days, truncated at a maximum window size of 15 days.

#### Regression techniques

We apply four different statistical approaches to quantify the impact of a NPI *M* on the reduction of *R*_*t*_ (see details in the SI).

#### Case-control analysis

The case-control analysis (CC) considers each single category (L2) or subcategory (L3) *M* separately and evaluates in a matched comparison the difference Δ*R*_*t*_ in the *R*_*t*_ between all countries that implemented *M* (cases) with those that did not implement it (controls) during the observation window. The matching is done on epidemic age and the time of implementing any response. The comparison is made via a linear regression model adjusting for (i) epidemic age (days after the country has reached 30 confirmed cases), (ii) the value of *R*_*t*_ before *M* takes effect, (iii) total population, (iv) population density, (v) the total number of NPIs implemented and (vi) number of NPIs implemented in the same category as *M*. With this design, we investigate the time delay of *τ* days between implementing *M* and observing Δ*R*_*t*_, as well as additional country-based covariates that quantify other dimensions of governance and human and economic development. Estimates for *R*_*t*_ are averaged over delays between 1 and 28 days.

#### Step function Lasso regression

In this approach, we assume that without any intervention, the reproduction factor is constant and deviations from this constant are caused by a delayed onset by *τ* days of each NPI on L2 (categories) of the hierarchical data set. We use a Lasso regularization approach combined with a meta parameter search to select a reduced set of NPIs that best describe the observed Δ*R*_*t*_. Estimates for the changes of Δ*R*_*t*_ attributable to NPI *M* are obtained from country-wise cross-validation.

#### Random forest regression

We perform a random forest (RF) regression, where the NPIs implemented in a country are used as predictors for *R*_*t*_, time-shifted *τ* days into the future. Here, *τ* accounts for the time delay between implementation and onset of the effect of a given NPI. Similar to the Lasso regression, the assumption underlying the random forest approach is that without changes in interventions, the effective reproduction number in a territory remains constant. But contrary to the two methods described above, the random forest represents a nonlinear model, meaning that the effects of individual NPIs on *R*_*t*_ do not need to add up linearly. The importance of a NPI is defined as the decline in the predictive performance of the random forest on unseen data if the data concerning that NPI is replaced by noise, also called permutation importance.

#### Transformers modeling

Transformers ^54^ have proven themselves as suitable models for dynamic discrete elements processes such as textual sequences due to their ability to recall past events. Here we extended the Transformer architecture to approach the continuous case of epidemic data by removing the probabilistic output layer with a linear combination of the Transformer output, whose input is identical to the one for the random forest regression, along with the values of *R*_*t*_. The best performing network (least mean squared error in country-wise cross-validation) is identified as a Transformer encoder having four hidden layers of 128 neurons, an embedding size of 128, eight heads, one output described by a linear output layer, and 47 inputs (corresponding to each category and *R*_*t*_). To quantify the impact of a measure *M* on *R*_*t*_, we use the trained Transformer as a predictive model and compare simulations without any measure (reference) to simulations where one measure is presented at a time to assess Δ*R*_*t*_. To reduce the effects of overfitting and multiplicity of local minima, we report results from an ensemble of Transformers trained to similar precision levels.

#### Estimation of the effective reproduction number

We use the R package EpiEstim ^55^ with a sliding time window of 7 days to estimate the time series of the effective reproduction number *R*_*t*_ for every country. We choose an uncertain serial interval following a probability distribution with a mean of 4.46 days and a standard deviation of 2.63 days ^56^.

#### Ranking of NPIs

For each of the four methods (CC, Lasso regression and TF), we rank the NPI categories in descending order according to their impact, i.e., the estimated degree to which they lower *R*_*t*_ or their feature importance (RF). To compare the rankings, we count how many of the 46 considered NPIs are classified as belonging to the top *X* ranked measures in all methods and test the null hypothesis that this overlap has been obtained from completely independent rankings. The *p*-value is then given by the complementary cumulative distribution function for a binomial experiment with 46 trials and success probability (*X/*46)^4^. We report the median *p*-value obtained over all *X* ≤ 10 to ensure that the results do not depend on where we impose the cutoff for the classes.

#### Co-implementation network

If there is a statistical tendency that a country implementing NPI *i* also implements NPI *j* later in time, we draw a directed link from *i* to *j*. Nodes are placed on the *y*-axis according to the average epidemic age at which the corresponding NPI is implemented; they are grouped on the *x* axis by their L1 theme. Node colours correspond to themes. The effectiveness scores for all NPIs are rescaled between zero and one for each method; node size is proportional to the rescaled scores, averaged over all methods.

#### Entropic country-level approach

Each territory can be characterised by its socio-economic conditions and the unique temporal sequence of NPIs adopted. To quantify the NPI effect, we measure the heterogeneity of the overall rank of a NPI amongst the countries that have taken that NPI. To compare countries which have implemented different numbers of NPIs, we consider the normalised rankings, where the ranking position is divided by the number of elements in the ranking list (i.e., the number of NPIs taken in a specific country). We then bin the interval [0, 1] of the normalised rankings into 10 subintervals and compute for each NPI the entropy of the distribution of occurrences of the NPI in the different normalised rankings per country:

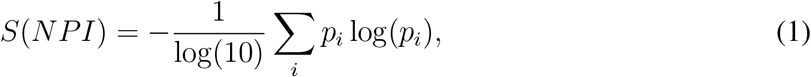

where *p*_*i*_ is the probability that the considered NPI appeared in the *i*-th bin in the normalised rankings of all countries. To assess the confidence of these entropic values, results are compared with expectations from a temporal reshuffling of the data. For each country, we keep the same NPIs adopted but reshuffle the timestamps of their adoption.

## Acknowledgements

We thank Alexandra Roux for her contribution to the coding of the interventions recorded in the data set used in this study. We thank David Garcia, Vito D.P. Servedio and David Hofmann for their contribution in the early stage of this work. NH would like to thank Luis Haug for helpful discussions. This work was funded by the Austrian Science Promotion Agency, FFG project under 857136, the WWTF under COV 20-001, COV 20-017 and MA16-045, the Medizinisch-Wissenschaftlichen Fonds des Bürgermeisters der Bundeshauptstadt Wien under CoVid004, and the project VET-Austria, a cooperation between the Austrian Federal Ministry of Social Affairs, Health, Care and Consumer Protection, the Austrian Agency for Health and Food Safety and the University of Veterinary Medicine Vienna.

## Author contributions

NH, LG, AL, VL, PK conceived and performed the analyses. VL, ST, PK supervised the study. ED contributed additional tools. NH, LG, AL, ADL, BP and PK wrote the first draft of the paper. ADL supervised the data collection on NPIs. All authors (NH, LG, AL, ED, ADL, VL, BP, ST, PK) discussed the results and contributed to the revision of the final manuscript.

## 6 SUPPLEMENTARY INFORMATION

### Case-control analysis

We perform a case-control regression analysis to quantify the impact of implementing an NPI measure on the effective reproduction number, *R*_*t*_. The central idea is to compare all countries that have implemented a certain measure with all countries that have not implemented this measure at the same stage of the epidemic while adjusting for several country-specific covariates through regression analysis. These covariates include timing (time-span between the day on which more than 30 cases were confirmed and the day the measure was implemented), baseline *R*_*t*_ (reproduction number before the measure was implemented), size and population density, as well as covariates for how many other measures have already been implemented. It is assumed that implementing a NPI on day *t* will have measurable impacts on *R*_*t*_ at day *t* + *τ*. We also study how various country-specific indicators for economic and human development as well as governance impact on these results.

### Exposure variable

We consider each NPI implemented in more than ten countries. We include measures published by ^25^ on two different resolution levels (L2 and L3) separately. Let *T*_*M*_ be the day on which a country implemented measure *M*. The covariates and outcome variables are measured relative to this point in time. As an exposure variable, we use a dummy variable, *X*, encoding whether a country has implemented the measure during the observation window or not.

**Figure S1:**
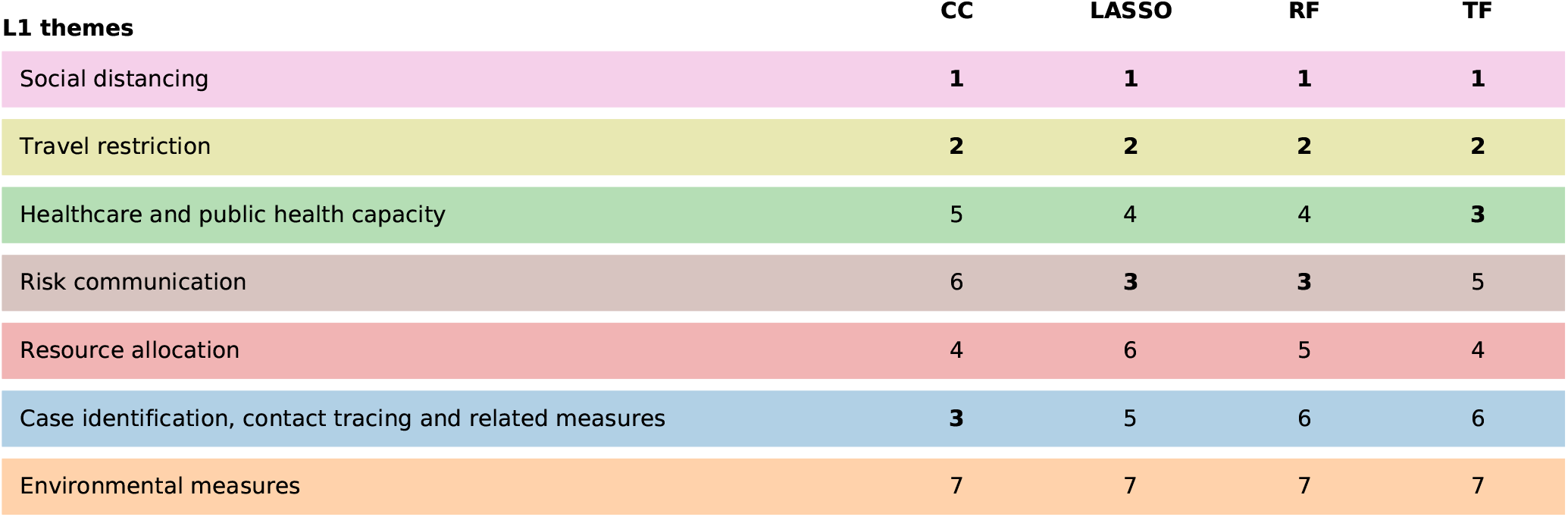
Comparison of effectiveness rankings on the coarsest hierarchical level for the case-control analysis (CC), LASSO regression (LASSO), random forest regression (RF), and transformer analysis (TF). To obtain a ranking of the eight different themes (L1) of NPIs, we sum the impacts of the 3 highest ranked categories of each theme and then rank the themes according to this cumulative impact. All methods indicate that NPIs belonging to the themes of social distancing, travel restrictions as well as healthcare and public health capacity lead to the most significant reductions of *R*_*t*_. Environmental measures (e.g., cleansing public places) are ranked least effective in each approach.

**Figure S2:**
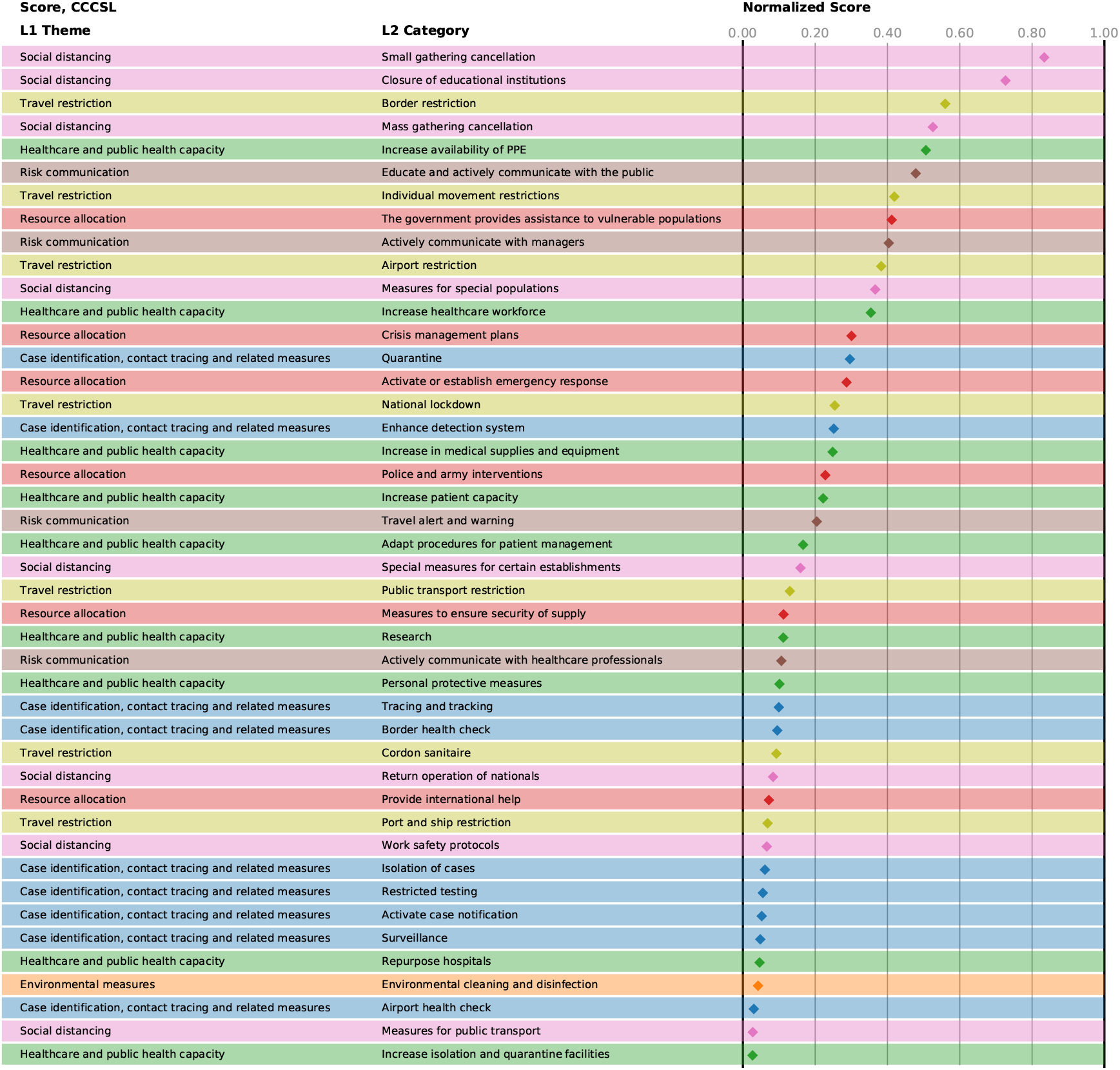
Normalised scores of the NPI categories in CCCSL, averaged over the four different approaches.

**Figure S3:**
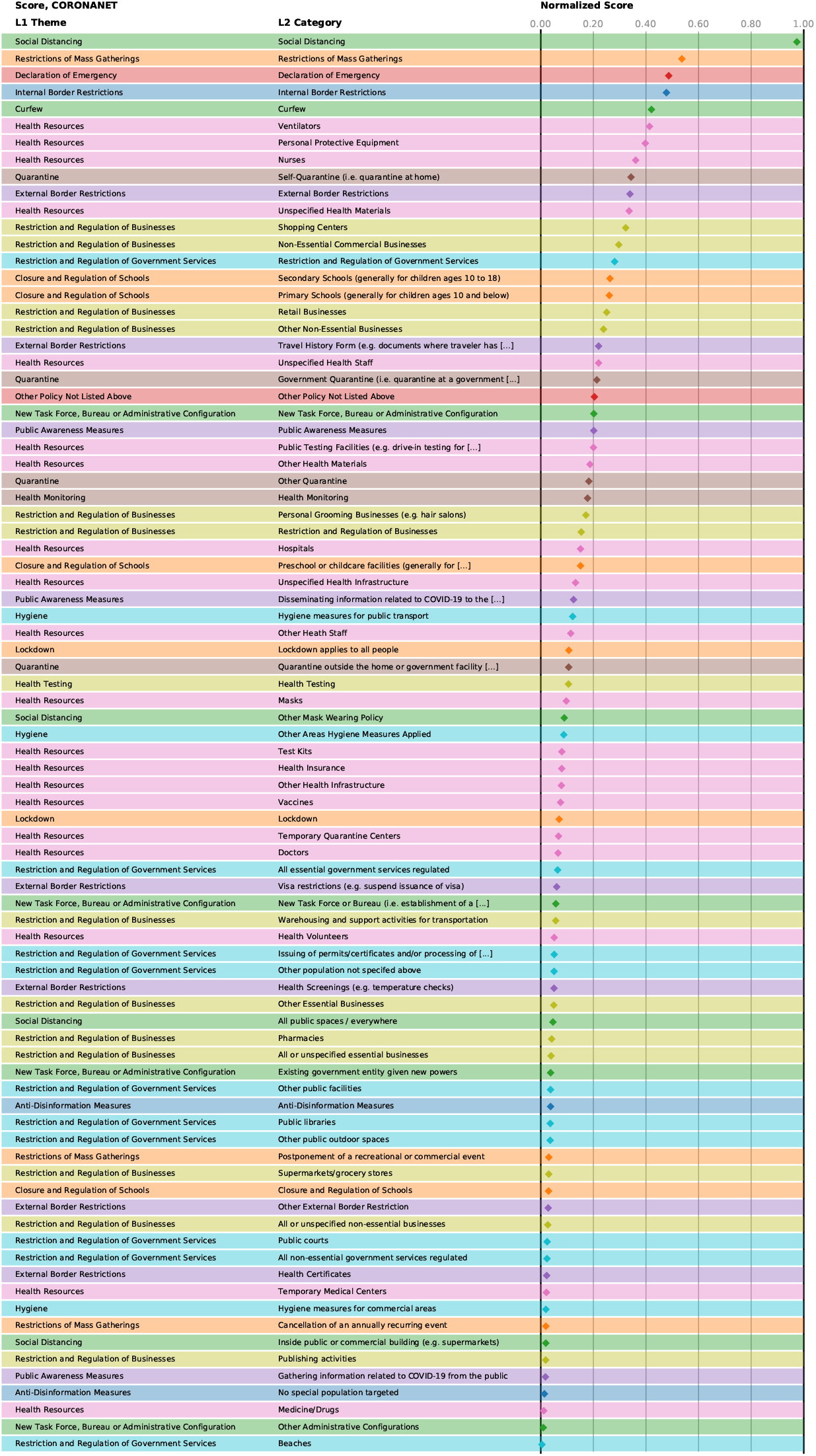
Normalised scores of the NPI categories in CoronaNet, averaged over the four different approaches.

**Figure S4:**
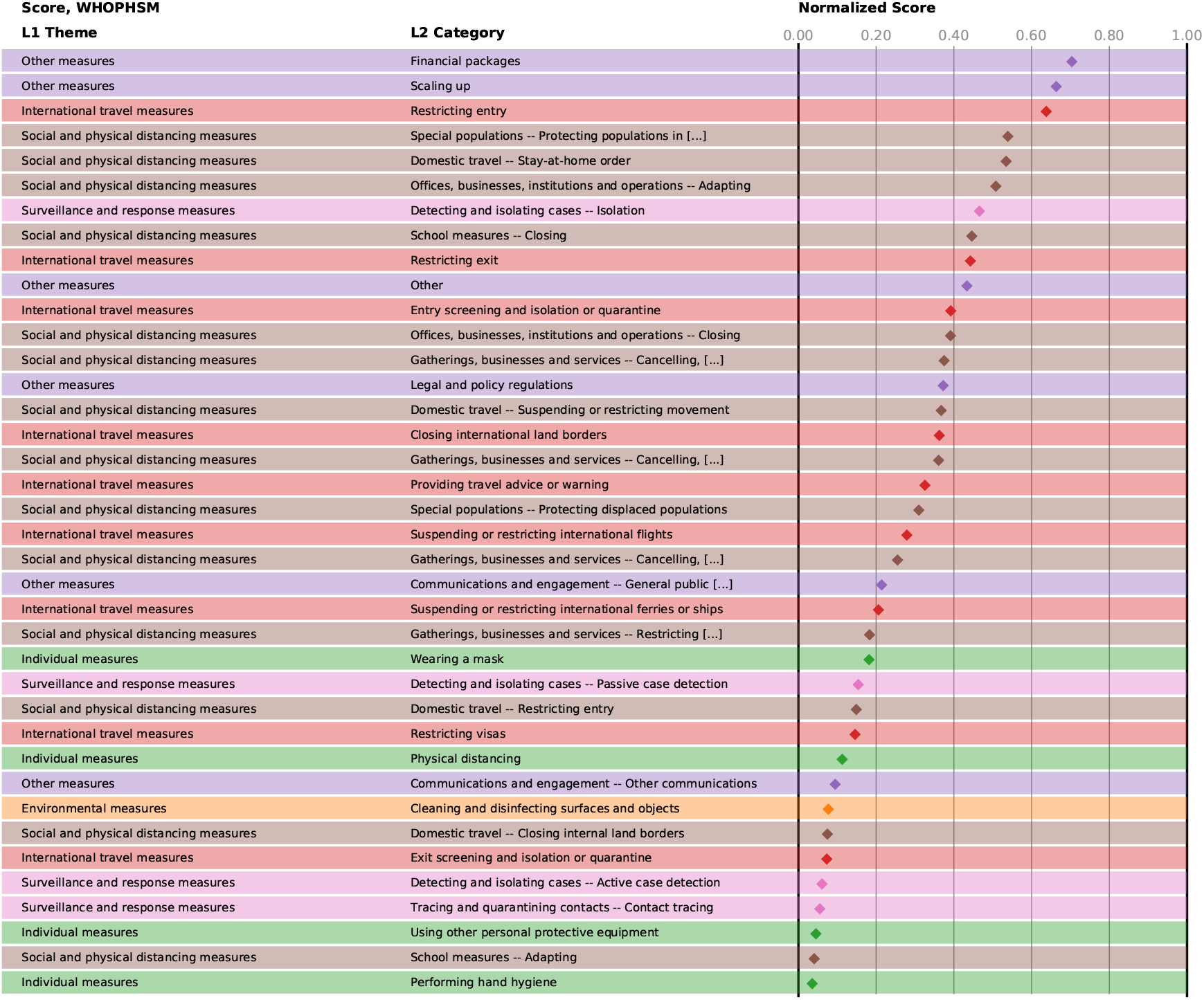
Normalised scores of the NPI categories in WHOPHSM, averaged over the four different approaches.

### Covariates

We include the following covariates in all analyses. First, a country’s epidemic age when measure *M* was implemented, *A*(*M*), defined as the number of days between the implementation of the measure, *T*_*M*_, and the first day with more than 30 confirmed cases, denoted by *T*_0_, giving *A*(*M*) = *T*_*M*_ − *T*_0_. Second, the baseline effective reproduction number, 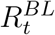, is taken at day *T*_*M*_ + *τ*. Third and fourth, we include for each country the logarithms of its total population *P* and population density *D* ^57^. Furthermore, we include the number *N*^*All*^(*T*_*M*_) of all implemented L2 measures and the number *N*^*L*2^(*T*_*M*_) of all measures (L2 or L3) from the same categories as *M* that have been implemented before *T*_*M*_. These covariates are supposed to capture whether a country introduces the intervention under consideration early or late in relation to its epidemic age and the number of measures already taken.

### Outcome variable

As a dependent variable in the regression, we consider the change in effective reproduction number after implementation of the measure, 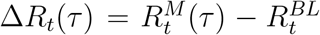, where 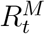 is the value of *R*_*t*_ taken on day *T*_*M*_ + *τ*. We report the average of Δ*R*_*t*_(*τ*) taken over time lags 0 < *τ* ≤ 28.

### Matching

As cases we include all countries implementing a given measure at a time *A*(*M*^*c*^) > *τ*. As controls we consider all other countries that have not implemented this measure within the observation window but that implemented any other intervention not more than one day sooner or later than its matched case. These matching criteria ensure that we do not introduce a bias by comparing countries that implemented any response with countries that did nothing at all and maybe had the epidemics already under control.

### Additional country variables

We consider variables that capture economic and human development, as well as different dimensions of governance. Economic development is measured by the country’s per capita GDP adjusted for power purchasing parity (PPP) ^58^. Human development is quantified by the human development index (HDI), an indicator provided by the United Nations Development Programme taking life expectancy, education, and per capita income into account ^59^. Finally, we consider six different dimensions of governance as quantified by the World Governance Indicators (WGI) ^60^. These indicators include Voice & Accountability (free media, the extent to which citizens participate in the government), Political Stability (including the absence of violence), Government Effectiveness (quality of public services), Regulatory Quality (the ability of government to implement sound policies), Rule of Law (the extent to which citizens abide by the rules of society) and Control of Corruption.

### Statistical analysis and multiple testing

A linear regression model of the form,

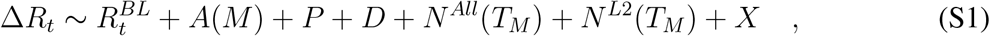

is evaluated for all NPIs meeting the inclusion criteria. We evaluate the above model for all measures and permissible delays *τ*. We adjust for multiple testing by controlling the false discovery rate at level 0.0001 via the Benjamini-Hochberg procedure. We investigate the impact of the additional country variables by evaluating models as in Eq. (S1) with an additional term for the considered indicator.

### Lasso regression

We assume that without any implemented NPIs, the *R*(*t*) is constant, and deviations are caused by a time-delayed onset of the effects of each NPI on L2. This hypothesis is an oversimplification of the reality, but it allows to estimate this time-delay itself and to quantify magnitudes of impacts for each NPI. A similar approach has been reported, for instance, in ^19^, although the authors use a smaller list of NPIs.

Formally, this model for a single territory *c* is captured in the expression

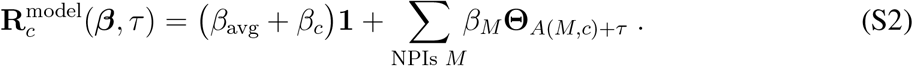

Here, bold symbols indicate vectors containing discrete time-series, starting at 30 confirmed cases for each country. The vector **Θ**_*t*_ models a discrete version of the Heaviside function with **Θ**_*t*_ = 0, …, 0, 1, …, 1, with the step from 0 to 1 at position *t*. In Eq. (S2), *A*(*M, c*) indicates the day when a NPI *M* is implemented in territory *c*, and *τ* is the offset in days to account for the time-delay until the NPIs affect the case numbers. The first term in Eq. (S2) is a constant composed of the average effects *β*_avg_ and potential territory-specific effects *β*_*c*_. For inference, we concatenate all territories into one large vector. Thus, in total we need to estimate 126/326/265 coefficients 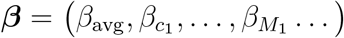 (1 constant, 79/218/224 territories and 46/107/40 NPIs for CCCSL/CORONANET/WHOPHSM) in this model.

Regression is carried out by minimizing the target function

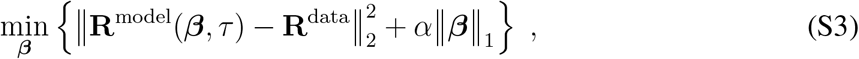

to obtain values for ***β***. The first term is the Residual Sum of Squares (RSS) of the model with respect to actual observation data. The second term penalises too large values of the coefficients ***β***, with *α* a penalty parameter that indicates the weight of this penalization in the optimization procedure. Here, ‖·‖_2_ and ‖·‖_1_ denote the 2-norm (euclidean distance) and 1-norm (sum of absolute values), respectively. The 1-norm for the penalizing term is characteristic for lasso regression ^61^, which acts as feature selection by estimating multiple coefficient values ***β*** as zero. Formally, this additional penalty term can be shown to be equivalent to assuming a Laplace-distribution prior on all values in ***β***. Thus, the tuning parameter *α* effectively balances the trade-off between two objectives: the goodness of fit to the data (RSS) with the complexity of the model (second term)^61, 62^.

### Cross-validation and meta-parameter search

This model includes two meta-parameters (*τ, α*), which are estimated by cross-validation to obtain a minimal RSS: At first, countries are randomly assigned to one of 10 groups. Then, each of these country groups is dropped from the vectors **R**, and coefficients ***β*** are estimated via the minimization in Eq. (S3). These coefficients ***β*** from the training group are used to compute an 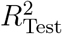 value of the model on the dropped countries to test how good the model can predict previously unseen observations. As the different country groups can contain a different number of observations, we compute the overall 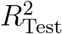 for a given set of (*τ, α*) as

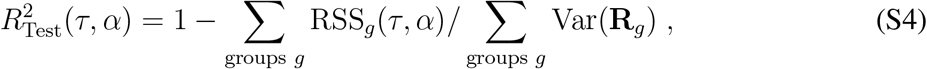

which weighs the individual coefficients of determination *r*^2^ for each test with the variance in the reproduction number **R**_*g*_ within the test group. This whole procedure is carried out on a grid of possible (*τ, α*) values, to find a set of meta-parameters, where the model Eq. (S2) can best describe the data. Fig. S5 depicts the resulting values of 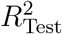 for a sufficient large range of meta-parameters. As the overall curve for 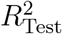 is relatively flat, we repeat the cross-validation 30 times with different assignments to the 10 country groups, which reduces the overall noise. The final 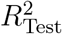 is then computed using the same weighting procedure in Eq. (S4) for the set of all 20 repetitions and each of the 10 territory groups within a repetition. We find that a time-offset of *τ* = 11*/*11*/*15 days and a penalty parameter *α* = 0.005623*/*0.00100*/*0.004217 are the best parameters to describe the observations with this model for the CCCSL/CORONANET/WHOPHSM, see Fig. S5.

### Final coefficient estimation

We estimate the ranges of each NPI effectiveness shown in Figs. S12, S16 and S20, using a territory-wise cross-validation, i.e., by reducing the sizes of test groups to one. For each estimation leaving out one territory, we compute the coefficients *β*_*M*_ for each NPI *M* and *β*_*c*_ for all territories *c*. We identify these coefficients *β*_*M*_ and *β*_*c*_ as the change Δ*R*_*t*_. Shown intervals contain 95% of all observed values, with the center indicating the median. From the overall 126/355/305 coefficients ***β***, the feature selection aspect of lasso regression finds 23/19/23 relevant NPI coefficients and 1/1/23 relevant territory coefficients, while other coefficients are estimated as zero (for CCCSL/CORONANET/WHOPHSM) Note that this does not indicate that these NPIs are useless to reduce the spread of the virus, but rather that the algorithm can explain all observations with a smaller number of coefficients. The number of selected coefficients is highly sensitive to the value of the penalty parameter, however, the more impactful NPIs are consistently present with similar coefficients (see Fig S5CFI).

**Figure S5:**
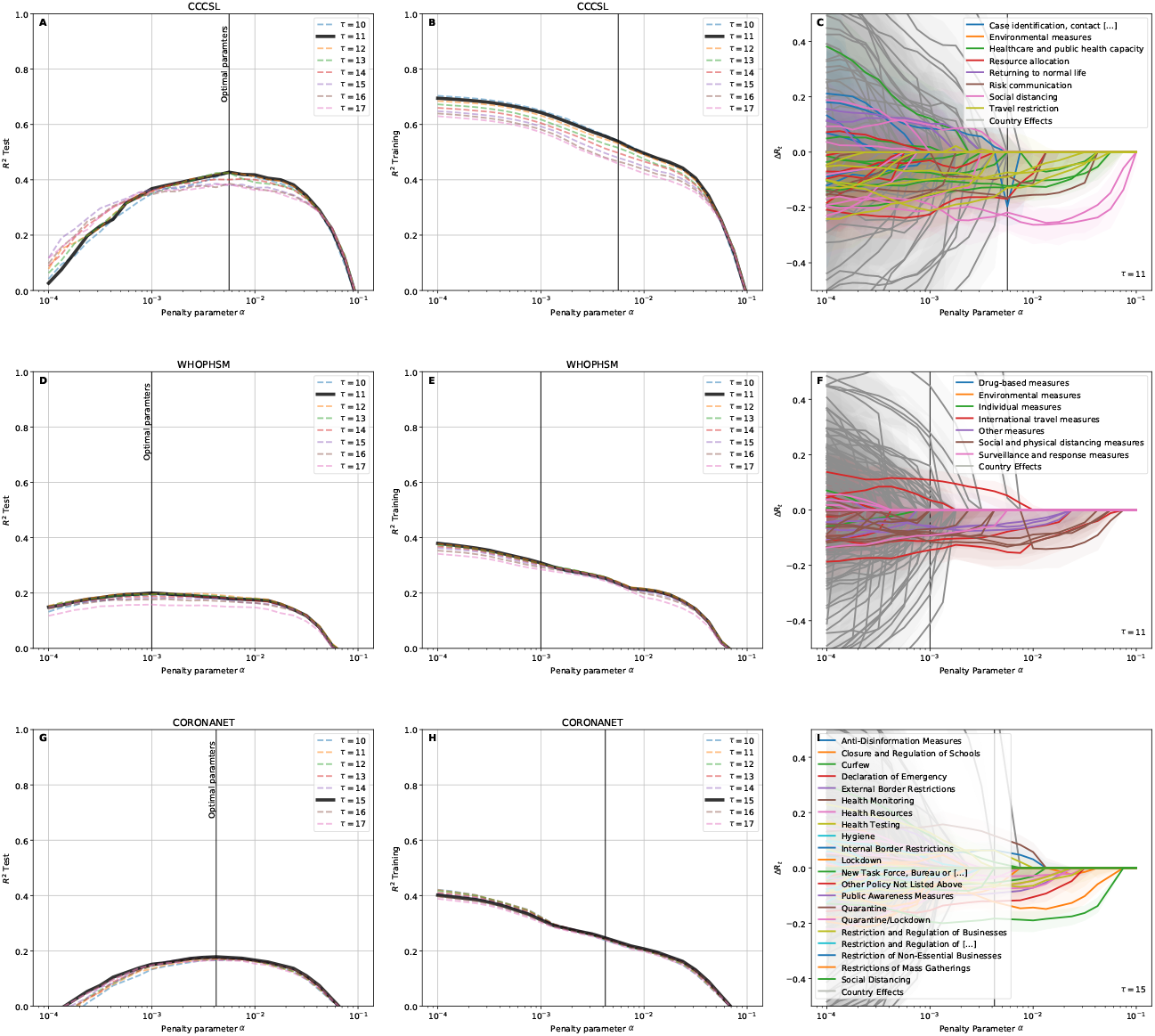
Cross-validation results for LASSO regression. (A,B,D,E,G,H) *r*^2^ values for the test and training data, averaged over 30 repetitions of assigning countries into 10 groups for each of the three data sets (CCCSL/CORONANET/WHOPHSM). With smaller penalty parameters *α*, 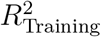 increases as the algorithm uses more NPIs to explain the training data. This overfitting, however, reduces the ability to fit the previously unseen test data. In contrast, for large penalty parameters *α*, not enough NPIs are used to explain the data, and the method cannot explain either training or test data. The optimal meta parameters are found as (*τ, α*)_CCCSL_ = (11, 0.005623), (*τ, α*)_CORONANET_ = (15, 0.004217) and (*τ, α*)_WHOPHSM_ = (11, 0.00100) (C,F,I) Effects of all NPIs and territories vary when changing the penalty parameter *α*. However, most of the effects of the significant NPIs (see Figs. S12, S16 and S20) stay roughly constant, only the number of significant NPIs increases.

### Random forest regression

We use random forest regression ^63^ as a third method to assess the impact of the implemented measures on the spreading of COVID-19, measured in terms of the effective reproduction number *R*_*t*_.

We represent for each country and each day of the observation period the NPIs which have been implemented in that country until that day in the form of a binary vector. The analysis is performed on the NPI categories (L2). The binary data on implemented NPIs in a given country on day *t* is regressed on the value of *R*_*t*_ in that country *τ* days later, *R*_*t*+*τ*_. The time shift *τ* accounts for the time delay between infection and case confirmation. We vary *τ* between *τ* = 0 and *τ* = 20.

Each random forest consists of 500 decision trees with maximum depth *d*. At each split of a node in one of the trees in the random forest, *m* randomly selected NPI categories out of the total number of 46 categories are considered. We employ bootstrapping, meaning that each decision tree is fitted on a different random subset containing 75% of the rows of the predictor matrix.

To implement the random forest regression, we use the RandomForestRegressor class from the python library scikit-learn ^64^.

### Cross validation

For each 0 ≤ *τ* ≤ 20, we use 10-fold country-wise cross validation to determine the optimal values of the maximum tree depth *d* and percentage of considered features *m*. We vary the parameters *d* and *m* in the range 1 ≤ *d* ≤ 15 and 1 ≤ *m* ≤ 100. For each combination of *d* and *m*, we randomly split the set of all territories into a training set and a test set. The random forest is trained on the training set data; subsequently, we measure the performance of this random forest in predicting the time series of *R*_*t*+*τ*_ for the countries in the test set. As a performance metric, we quantify the difference between the predicted time series of *R*_*t*+*τ*_ and the observed one by using the coefficient of determination *r*^2^. We repeat the same procedure 10 times with different random splits of the set of territories into training and test set. Then we take the mean of the coefficient of determination over the 10 splits to obtain the average out-of-sample performance of the random forest for this combination of *d* and *m*. The heatmap in Fig. S6 shows the dependence of the performance of the random forest on unseen data depending on the parameters *d* and *m* for fixed time shift *τ* = 10.

### Feature importance

To quantify the importance of a NPI, we measure the loss of predictive performance of the random forest if the information carried by the given NPI is replaced by noise. This measure of feature importance is also known as permutation importance. If the loss in performance is high for a given NPI, then we conclude that knowledge of the implementation status of this NPI is important for predicting *R*_*t*+*τ*_.

Specifically, for each time shift 0 ≤ *τ* ≤ 20 and each NPI *M*, we measure the reduction in performance of the random forest in predicting *R*_*t*+*τ*_ for unseen data when the values in the column corresponding to *M* are randomly shuffled. The maximum tree depth *d* and the percentage *m* of features considered are set to their optimal values as determined in the cross-validation step. To obtain sharp estimates for the permutation importance of the different NPIs we use repeated 10-fold country-wise cross validation with 100 repetitions. Different NPIs attain their maximum value for different values of *τ*.

**Figure S6:**
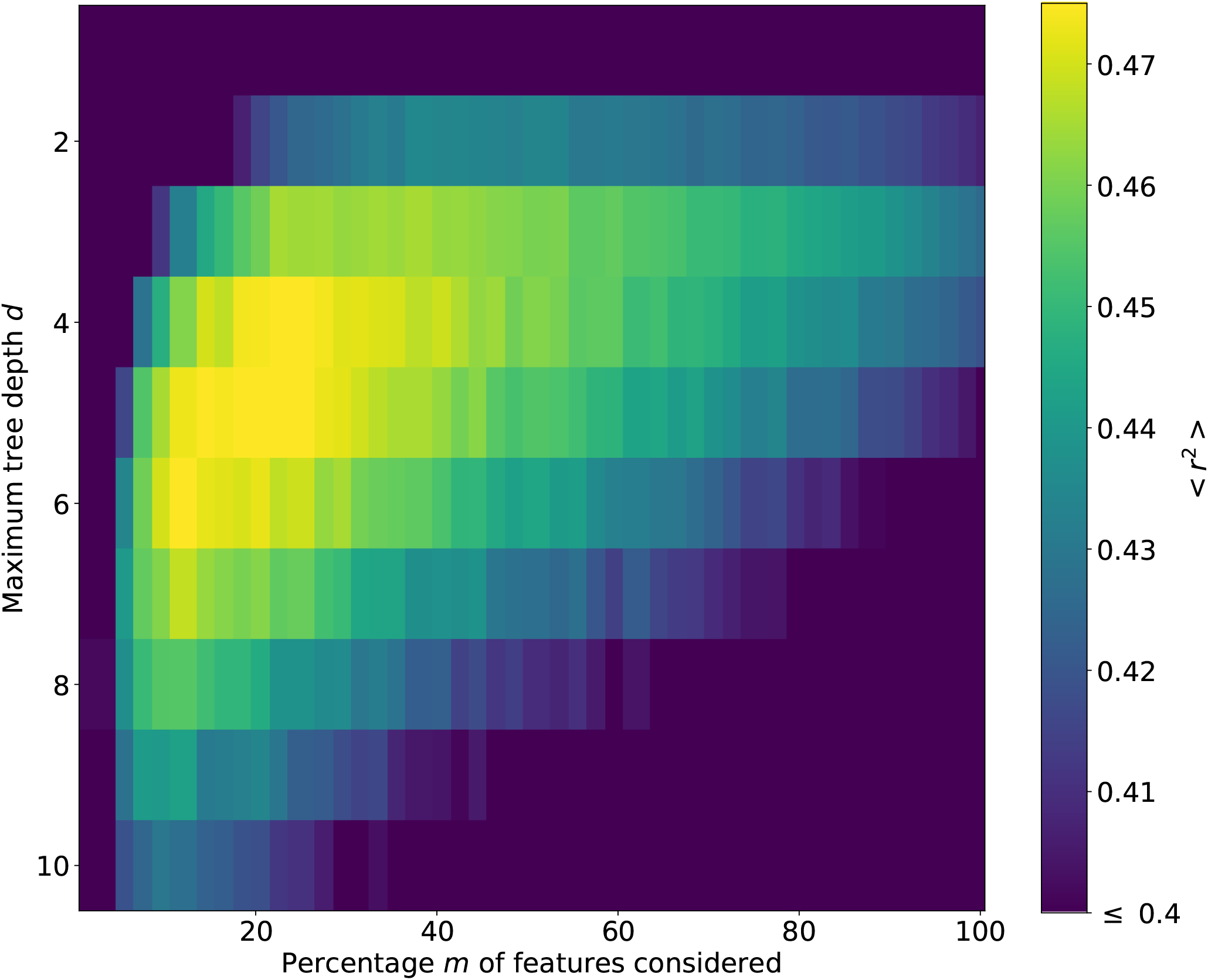
Heatmap of the predictive performance of the random forest depending on the maximum tree depth *d* and the percentage of features *m* considered at each split for time shift *τ* = 10. The predictive performance is measured in terms of the coefficient of determination (*r*^2^) on unseen data, averaged over 10 random splits of the territories into training and test sets.

### Transformers modeling

The temporal and dynamic nature of epidemic propagation finds a suitable tool to model such a temporal evolution in Transformers ^54^. This neural network can recall past events presented at the input by leveraging its innate ability to take into account the previous information. The intrinsic Transformer architecture ensures this ability for all the temporal data is represented and considered at once without recurrent connections as in Recurrent Neural Networks. At a generic time *t*, to approximate the value of the effective reproduction number at time *t* + 1, *R*_*t*+1_, we use a Transformer whose input is the daily representation of the adopted measures in a given country in binary form, similar to the encoding performed in Random Forest, along with the value of *R*_*t*_ measured in the same day. The best performing network has been identified as having four hidden layers of 128 neurons, an embedding size of 128, 8 heads, one output described by a linear output layer, and 54 inputs (*measures* ⊕ *R*_*t*_, where ⊕ is the concatenation operator). NPIs are represented through a theta function over time, whose step position corresponds to the day when the specific measure has been adopted in the selected country. For the neural network training, 10 out of 78 territories were chosen as the validation set, and the best neural network was found when the mean square error between the values of the validation set and the corresponding network outputs reached the minimum amount. Different Transformers training can lead to slightly different predictions. Although the main trends of forecast maintain a substantial similarity across the various training, the corresponding time evolution may change when some containment measures are perturbed. This issue is possibly due to a large number of local minima affecting the neural network’s energetic panorama. To address this problem and to provide an adequate assessment of the importance of the measures, we built an ensemble of trained Transformers with a comparable level of training precision.

### Transformer ranking assessment

This approach has been chosen to be consistent with the other methods implemented in this paper, in which the impact of the NPIs is related to the comparison between countries having or not having implemented a measure. Therefore the comparison is performed between the normal prediction of the Transformer after the application of a given NPI and the corresponding prediction when the same measure is removed. The difference between the predictions quantifies the impact of that NPI on the overall behaviour of *R*_*t*_ of the selected country. The final relative importance of the containment measure *meas*_*i*_ is then given by 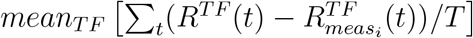, where *mean*_*T F*_ (·) is the mean operator over the RNN ensemble, *R*^*T F*^ (*t*) is the reproduction number predicted by the *TF* th network, 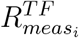*(t)* is the corresponding temporal prediction when the measure *meas*_*i*_ is removed, and *T* is the period over which the assessment is performed.

### Transformer model at country level

Implementing a Transformer as a general model for describing the dependence on the measures of *R*_*t*_ allows, at the same time, to evaluate the impact of the measures on a single country. Each country is identified by a specific temporal sequence of applied measures and a unique *R*_*t*_ evolution over time. Then, we can interrogate the Transformer to assess the importance of every single measure for that particular context by removing the measure under investigation from the specific sequence as in the general knockout case and building an associated ranking of NPIs for each country. For example, Fig. S7 shows the impact on *R*_*t*_ of the NPI “Small gathering cancellation” in Italy when such a measure is removed from the temporal sequence of adopted NPIs. This approach allows us to assess the country-specific NPI ranking. By comparing these ranking across all countries, we measure the specificity of the impact of its measures.

### What-If experiments with Transformers

As mentioned above, Transformers offer the possibility to perform what-if experiments, i.e., exploring different scenarios corresponding to time-series of events different from the actual ones. This approach opens the possibility to examine scenarios corresponding to different combinations of NPIs. In the specific case of the Non-Pharmaceutical Interventions to mitigate the spread of COVID-19, starting from the actual sequence of NPIs adopted by one particular country, we tested the efficacy of artificial sequences by removing specific NPIs or shifting their adoption to other days. In this framework, one can compare the actual evolution obtained through the synthetic sequence of actions to suitable reference sequences. More specifically, we adopted the Transformers to assess what would have happened if a given NPI had been adopted on a different day with respect to the actual day of adoption. To this end, we adopted the following procedure:

- for each country and each NPI, we first compute a *knockout evolution* of the system by letting the Transformer simulate the evolution of the system once the specific NPI has been removed.
- we create a synthetic sequence by keeping the sequence of NPIs of the country untouched except for the specific NPI that is positioned in a generic day *t*_*i*_. For that sequence, we compute the evolution of the system as given by the Transformer, i.e., we calculate the time evolution of *R*_*t*_ for the specific synthetic sequence.
- We repeat the above operation for a generic choice of the day of adoption, *t*_*i*_, of the specific NPI.
- For each country and each NPI, we compute the average difference between the synthetic evolution of *R*_*t*_ with the specific NPI positioned at *t*_*i*_ and the knockout evolution over the 30 days following *t*_*i*_. The average here is performed over several realisations of the Transformers.
- The outcome is a series of curves that, for each NPI, report the variation of *R*_*t*_, Δ*R*_*t*_, averaged over all the countries that adopted that NPI and over several realisations of the Transformers.

**Figure S7:**
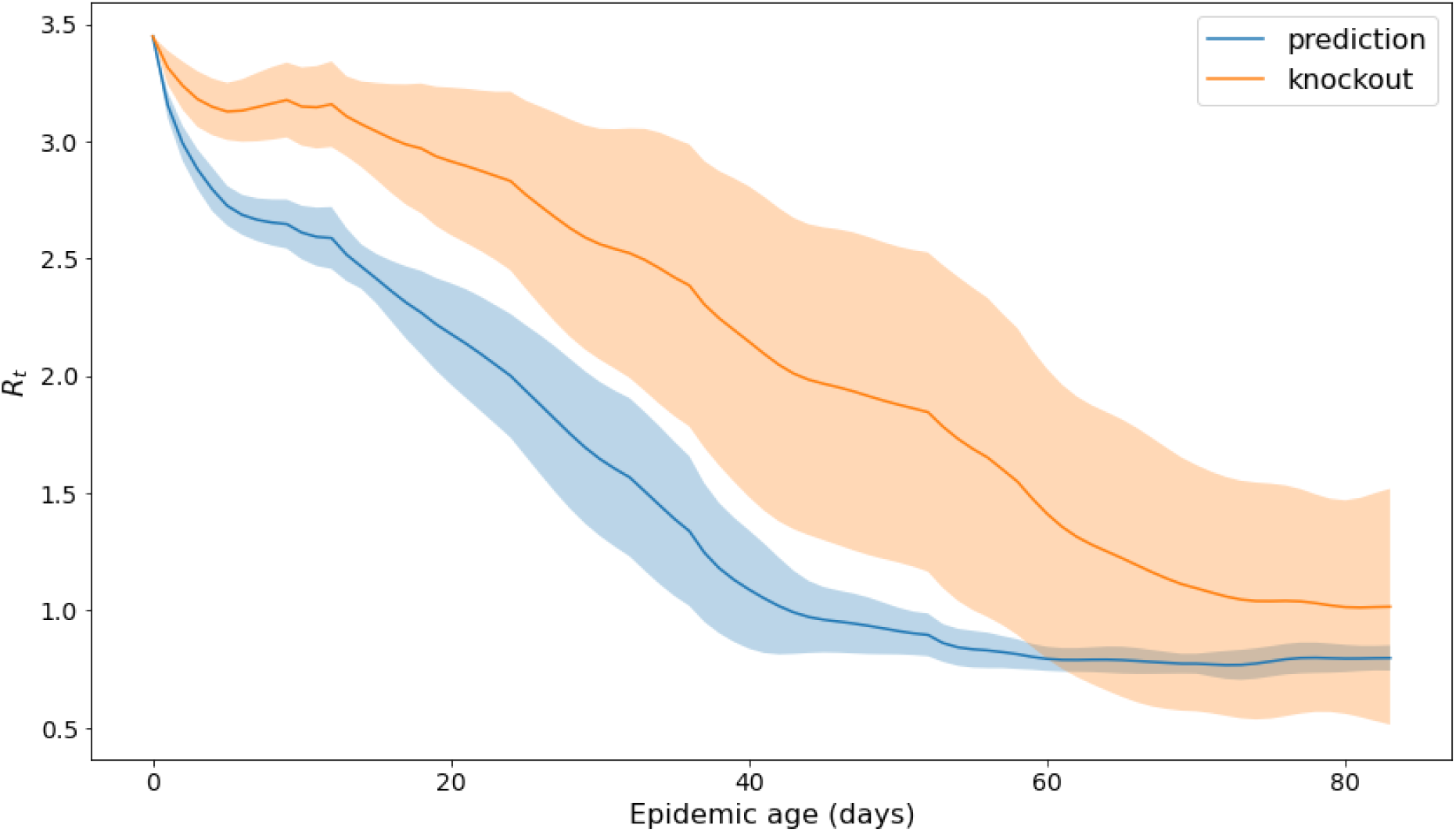
Example of the country-specific impact of an NPI. The plot shows the *R*_*t*_ increase when the NPI “Small gathering cancellation” is removed from the temporal measures sequence in Italy. Shaded areas are the standard deviations of the Transformer prediction averaged on the networks ensemble.

Fig. S8 shows the effect of the selected NPI “Small gathering cancellation” in Italy if it had been adopted on different days, compared to the case where the same NPI is absent (knockout evolution). The reduction of *R*_*t*_ is visible, while the overall impact decreases as one shifts the day of adoption ahead. The Transformer evolution, when the selected NPI is adopted on any other day, takes into account the effect of all the other NPIs adopted in Italy and kept fixed throughout the simulation.

Fig. S9 reports the evolution of Δ*R*_*t*_ for a selection of NPIs that display a “the earlier, the better” behaviour, that is, whose ability to reduce *R*_*t*_ tends to decrease with the epidemic age of their adoption.

**Figure S8:**
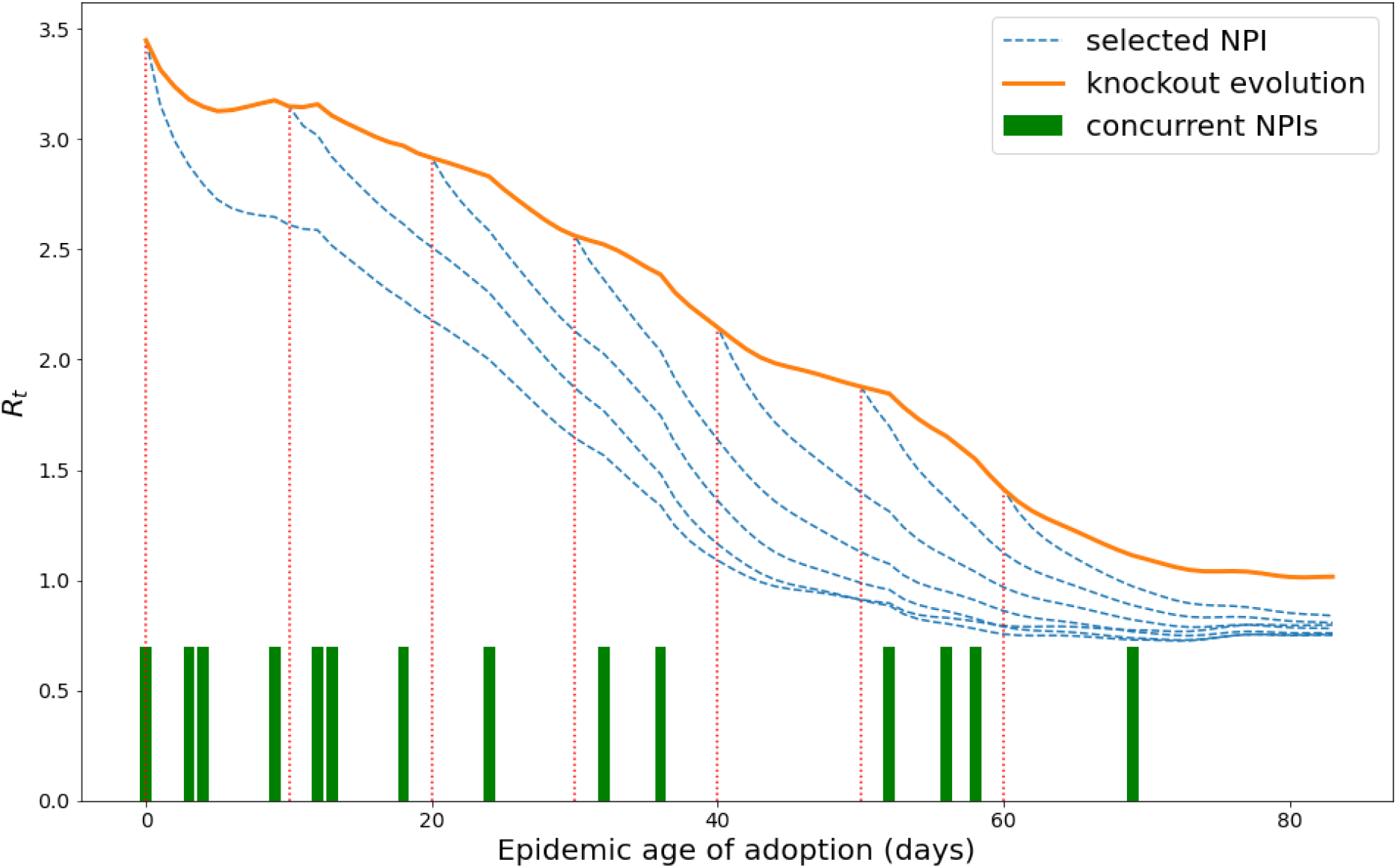
Example illustrating the what-if experiment described in the text. Here we consider the NPI “Small gathering cancellation” in Italy and we simulate what would happen if it had been taken at epidemic ages 0, 10, 20, 30, 40, 50, 60. We then compare this evolution with an evolution obtained through a knowknout of the same NPI (knockout evolution). The other concurrent NPIs and their days of adoption are kept fixed throughout the simulation (green rectangles at the bottom).

**Figure S9:**
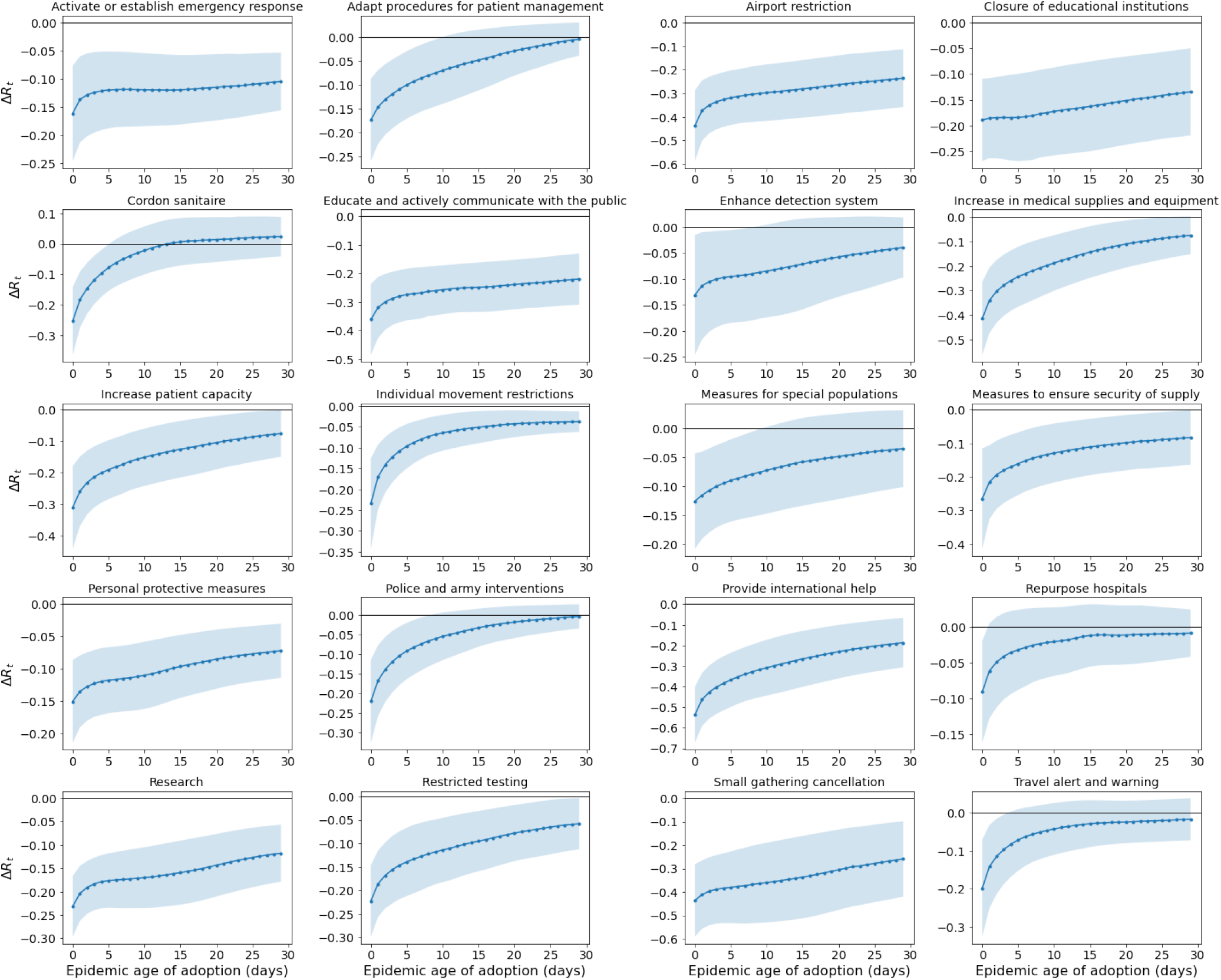
Outcome of the what-if experiment performed with the Transformers. We report, for several NPIs, the behaviour of the averaged variation of *R*_*t*_, Δ*R*_*t*_ (see text for the definition), when the NPI is adopted at a generic epidemic age compared with an evolution in which that NPI has been knocked-out. Negative (Positive) values of Δ*R*_*t*_ indicate a decrease (increase) of *R*_*t*_.

### Results from the Case-control Analysis on country covariate effects

The impact of different country variables, including demographic and governance indicators, as well as measures for human and economic development, are shown in Fig. S10. This heatmap presents the average standardised coefficient values from all regression models for a given NPI category (L2) that yield a significant impact on *R*(*t*) for various delays (the average is taken over the delays). The standardised coefficient values are the coefficient values (estimated change in *R*_*t*_) divided by its standard error (SE); the *t*-Statistic. The significant NPIs show effect sizes in reducing *R*_*t*_ of about 5 standard errors. Note that no statements concerning causal relationships can be made based on these findings; we report correlations from a cross-sectional analysis.

A number of NPIs appear to be less effective in countries with a high population density and low GDP ^65^. Human development, as quantified by the Human Development Indicator, has no or only very mild impacts on measure effectiveness. The governance indicators Political Stability and Voice & Accountability are negatively correlated with the effectiveness of most measures. Most of the remaining governance indicators show either no or moderately negative correlations. There are, however, exceptions for some interventions that show consistently positive correlations with government effectiveness, regulatory quality, rule of law and control of corruption, in particular measures to increase the availability of medical supplies and PPE. It is unlikely that these results are confounded by poor reporting in less developed countries, as such biases would affect *R*_*t*_ values from before and after measure implementation similarly and hence cancel each other out. Although there is no clear pattern concerning the influence of the total number of NPIs already implemented, there seems to be a trend regarding the number of NPIs of the same category. As a general tendency, having already implemented measures from a given category makes additional NPIs from the same category more effective, hinting at a relationship where one measure (e.g., closure of restaurants) amplifies the effectiveness of other related measures (e.g., home office).

### Results from the Random forest regression

Figs. S13, S17 and S21 give the rankings of the different NPIs in descending order according to their feature importance for the three analyzed datasets.

### Results from Transformers Analysis

We trained 20 different Transformers after selecting the best out of 100 training procedures to build a representative ensemble of networks to address multiple local minima issues. All the Transformers reached similar values of test loss with a relative deviation of less than 5%. The NPIs impact evaluation is performed by averaging the difference between the prediction and the prediction when a given measure is removed. The area under such a difference normalized by the temporal amplitude represents the mean variation of Δ*R*(*t*).

## 7 Robustness check and validation

We check the robustness of the results obtained against removal of the Americas (North and South America), Asia+Oceania, and Europe+Africa in the CCCSL dataset — see Figs. S24–S26. Note that the only African countries in the data set are Ghana, Senegal and Mauritius, and the single Oceanian country is New Zealand. Table S1 shows the number of consensus measures (i.e. NPIs with a significant effect in all methods), as well as the expected number of consensus measures one would expect under the assumption that the significant measures would be distributed independently for the different methods.

**Figure S10:**
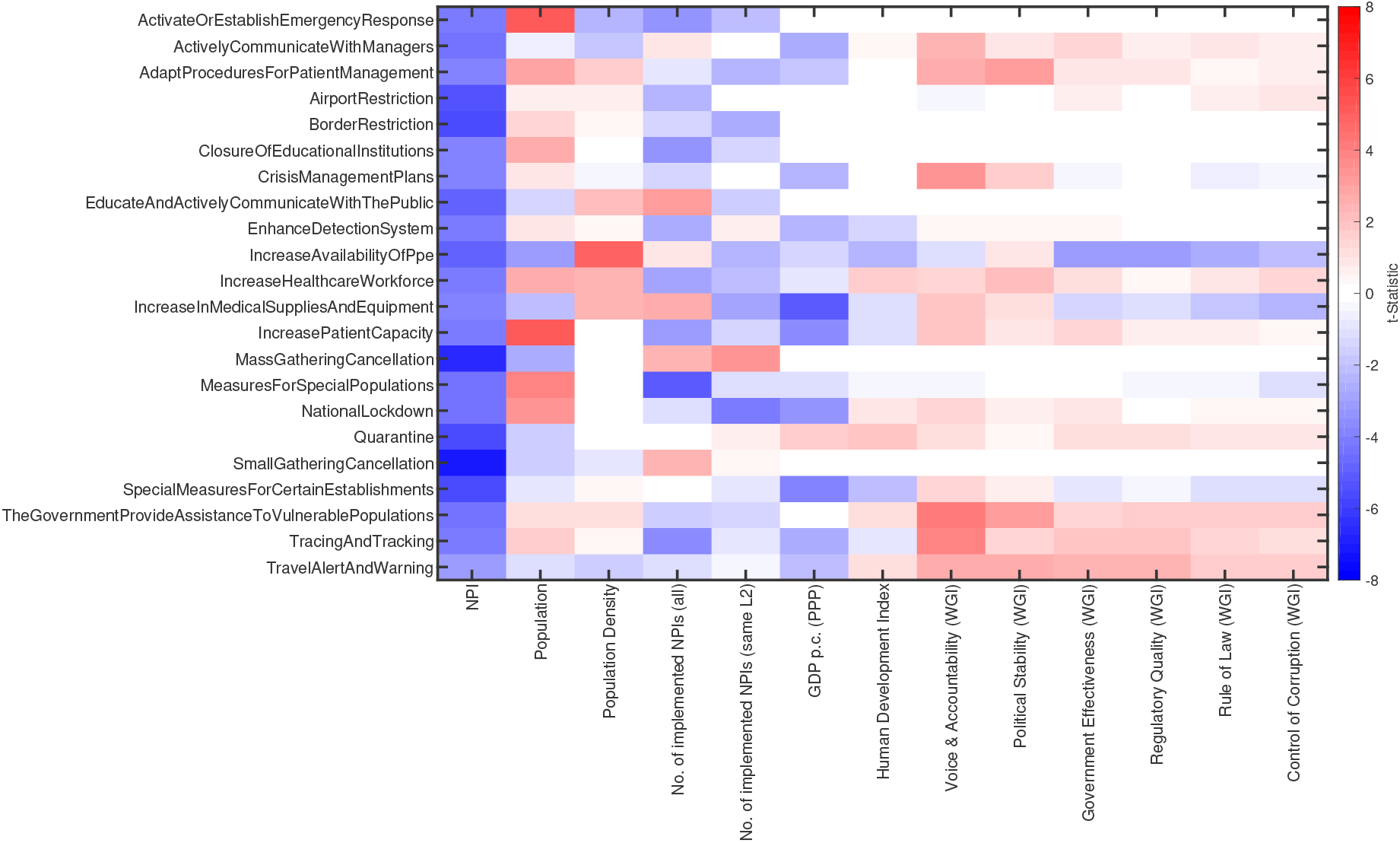
Impacts on *R*(*t*) of country variables on measure effectiveness. The heatmap gives the average effect size (t-statistic) for a given NPI category (L2) (rows) and a country variable (columns). Blue (red) color indicates that the variable is positively (negatively) correlated with measure effectiveness.

**Figure S11:**
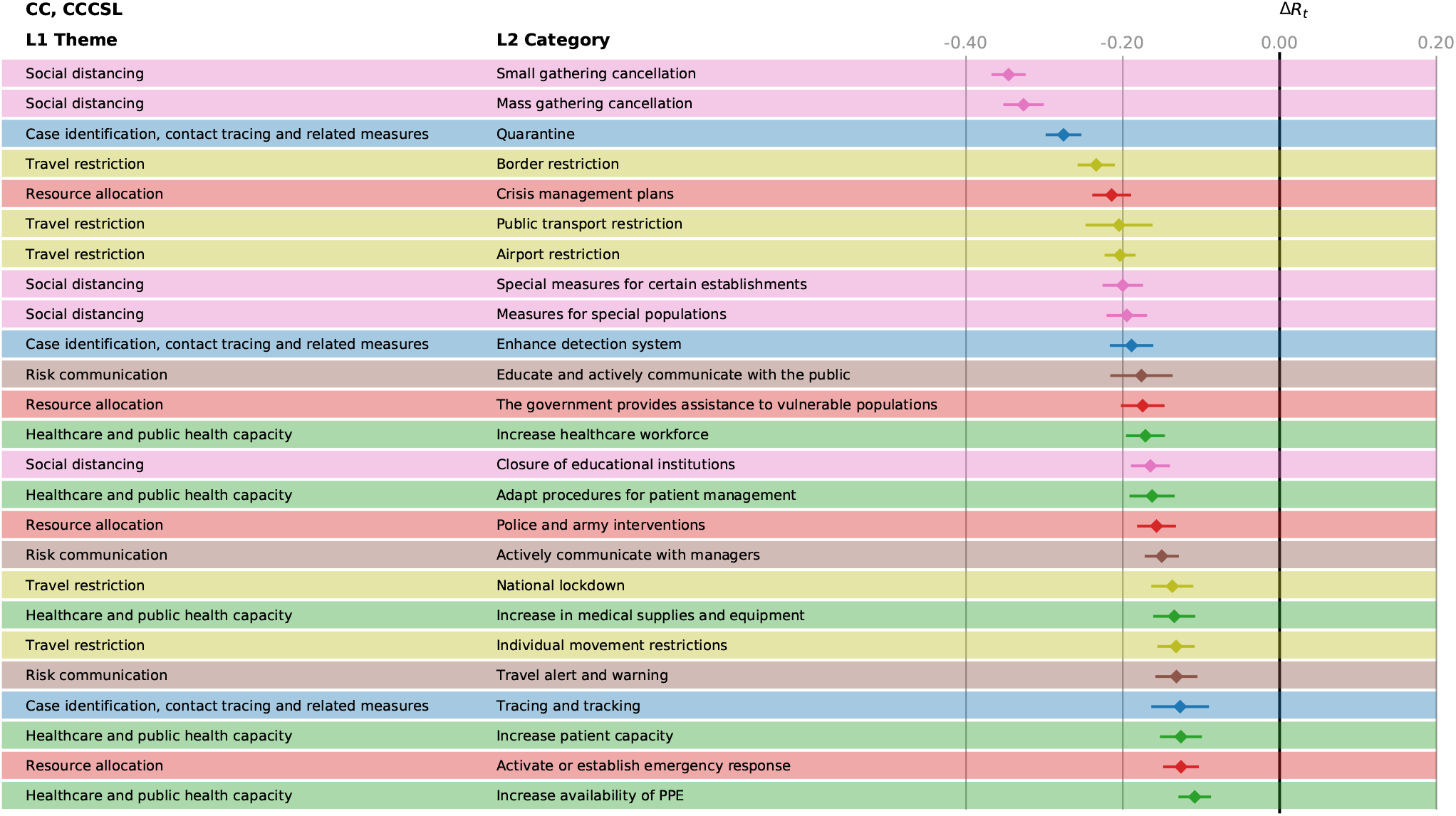
Effectiveness, Δ*R*_*t*_ of the different NPIs in the CC analysis for CCCSL. The horizontal bars mark the 95% confidence intervals.

**Figure S12:**
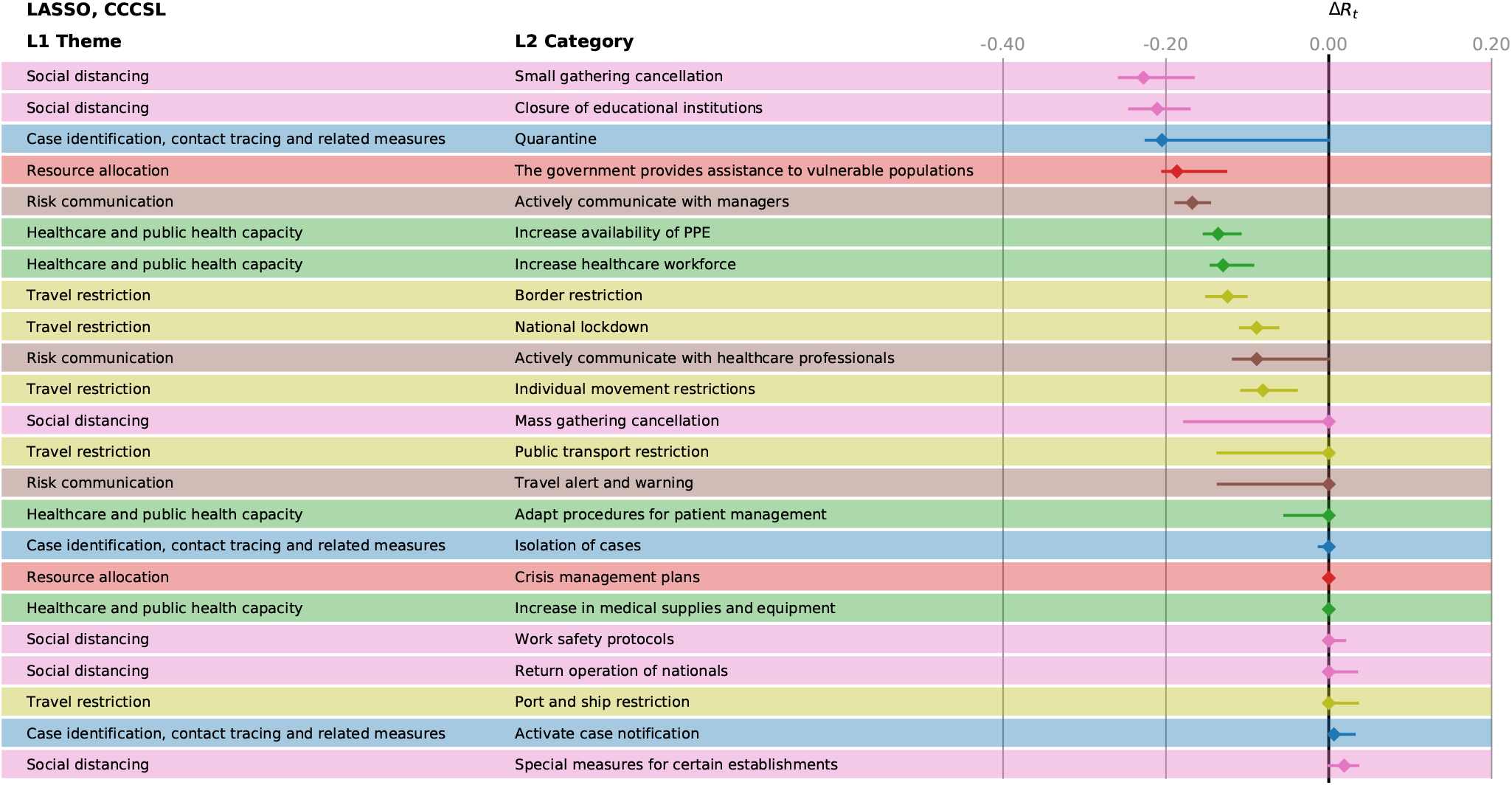
Effectiveness, Δ*R*_*t*_ of the different NPIs in the LASSO analysis for CCCSL. The horizontal bars mark the 95% confidence intervals.

**Figure S13:**
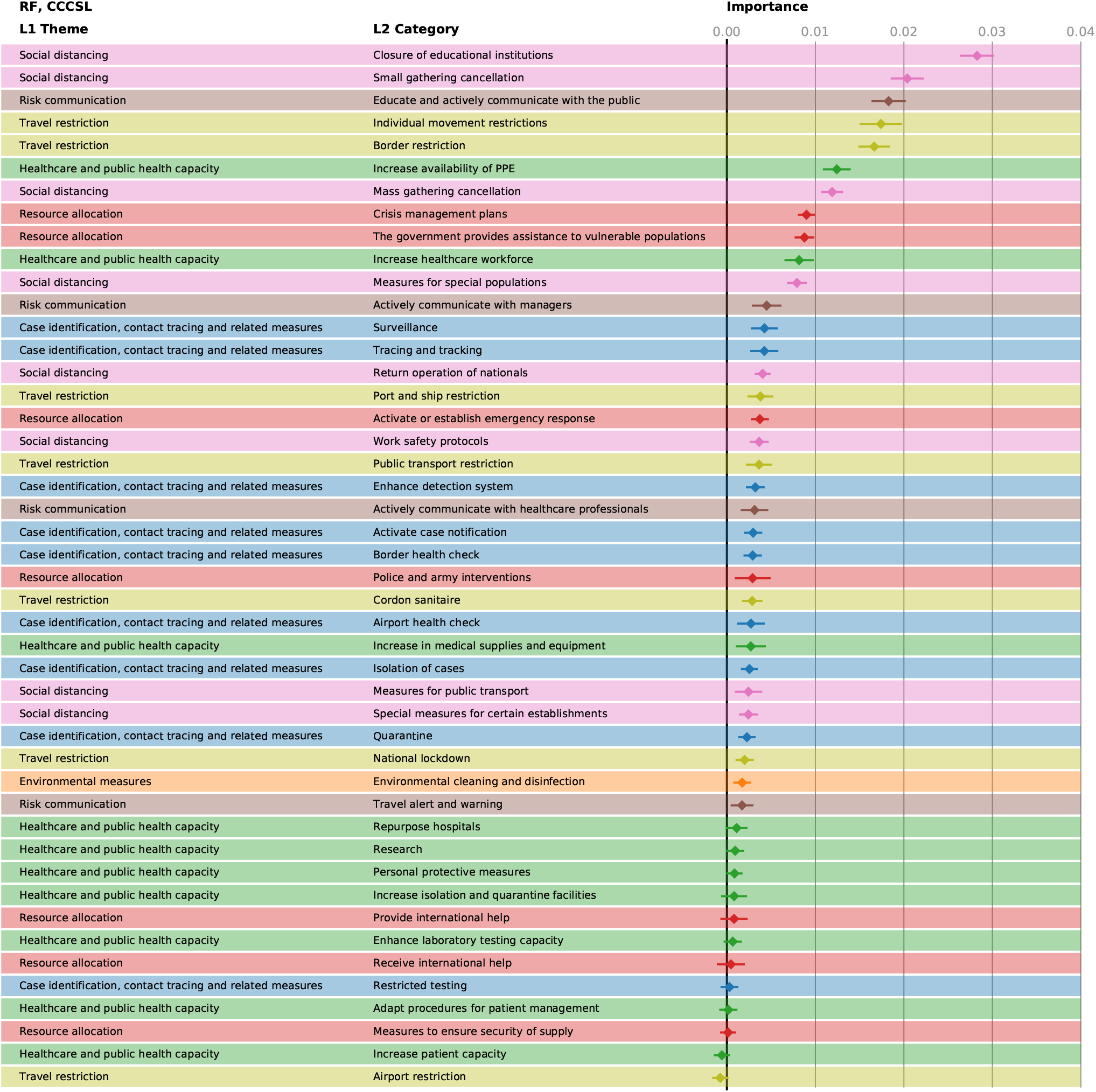
Feature importance of the different NPIs in the random forest model for CCCSL. The horizontal bars mark the 95% confidence intervals.

**Figure S14:**
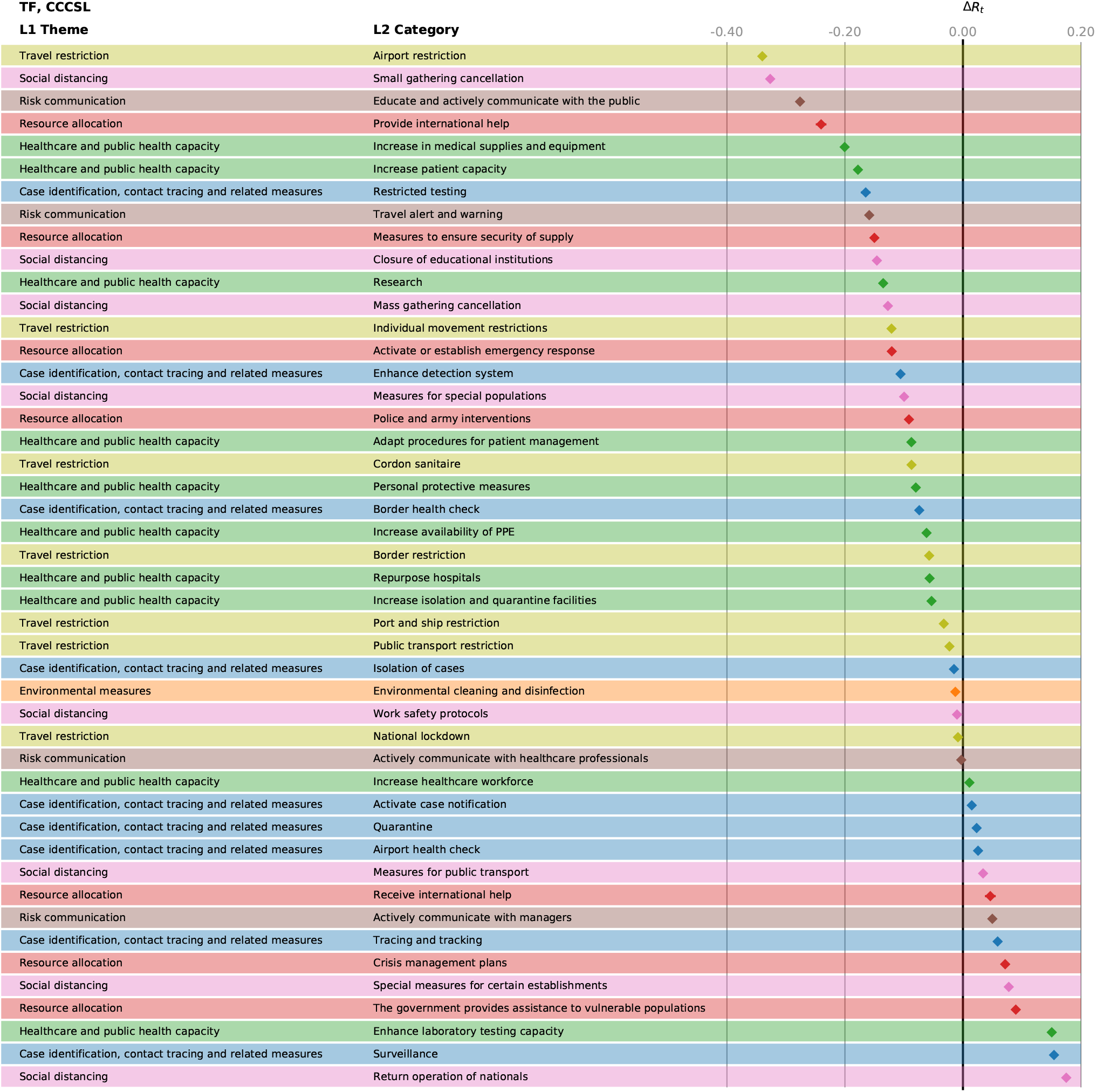
Effectiveness, Δ*R*_*t*_ of the different NPIs in the Transformer analysis for CCCSL. The bars marking the 95% confidence intervals are too small to be visible.

**Figure S15:**
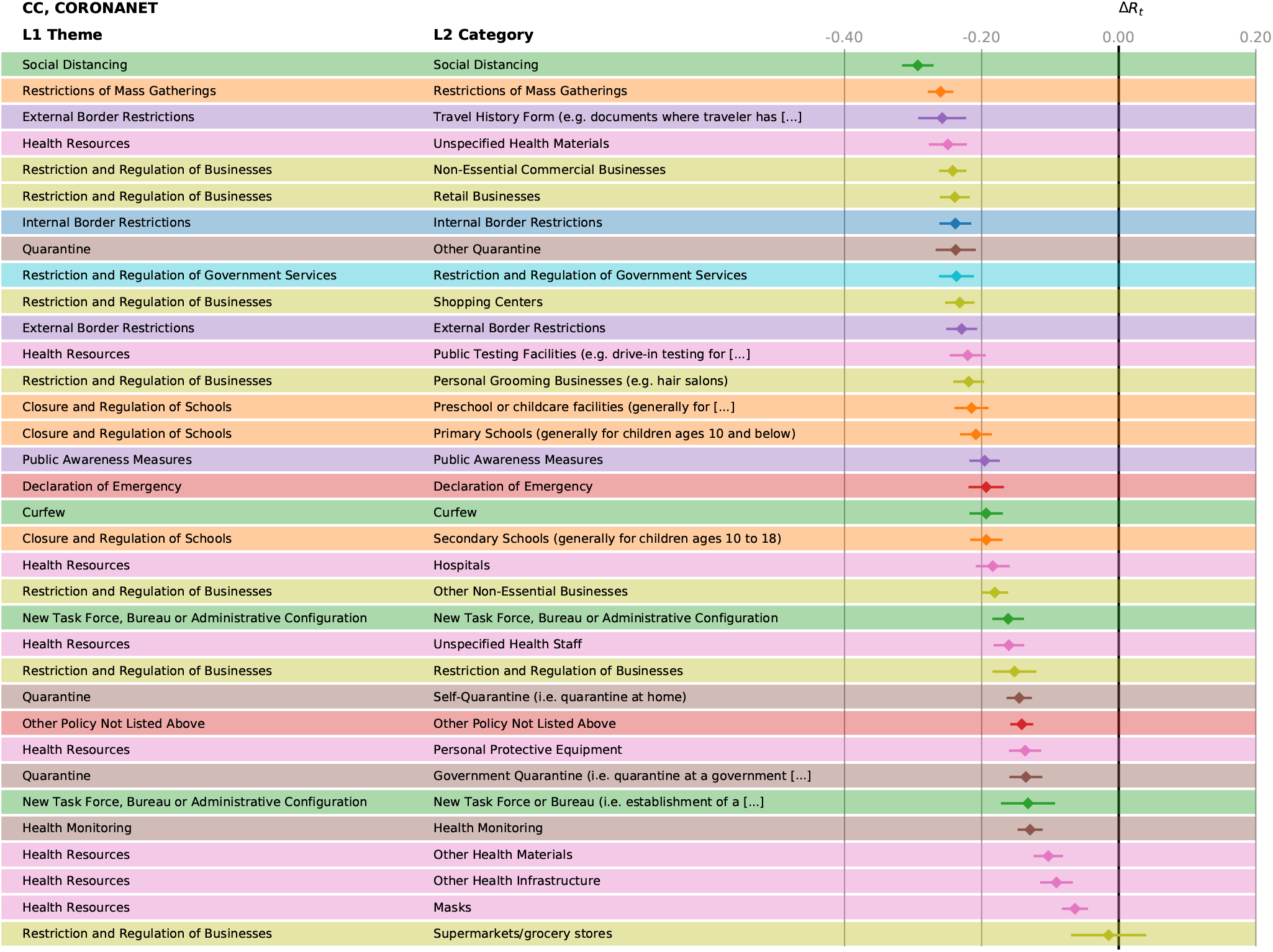
Effectiveness, Δ*R*_*t*_ of the different NPIs in the CC analysis for CORONANET. The horizontal bars mark the 95% confidence intervals.

**Figure S16:**
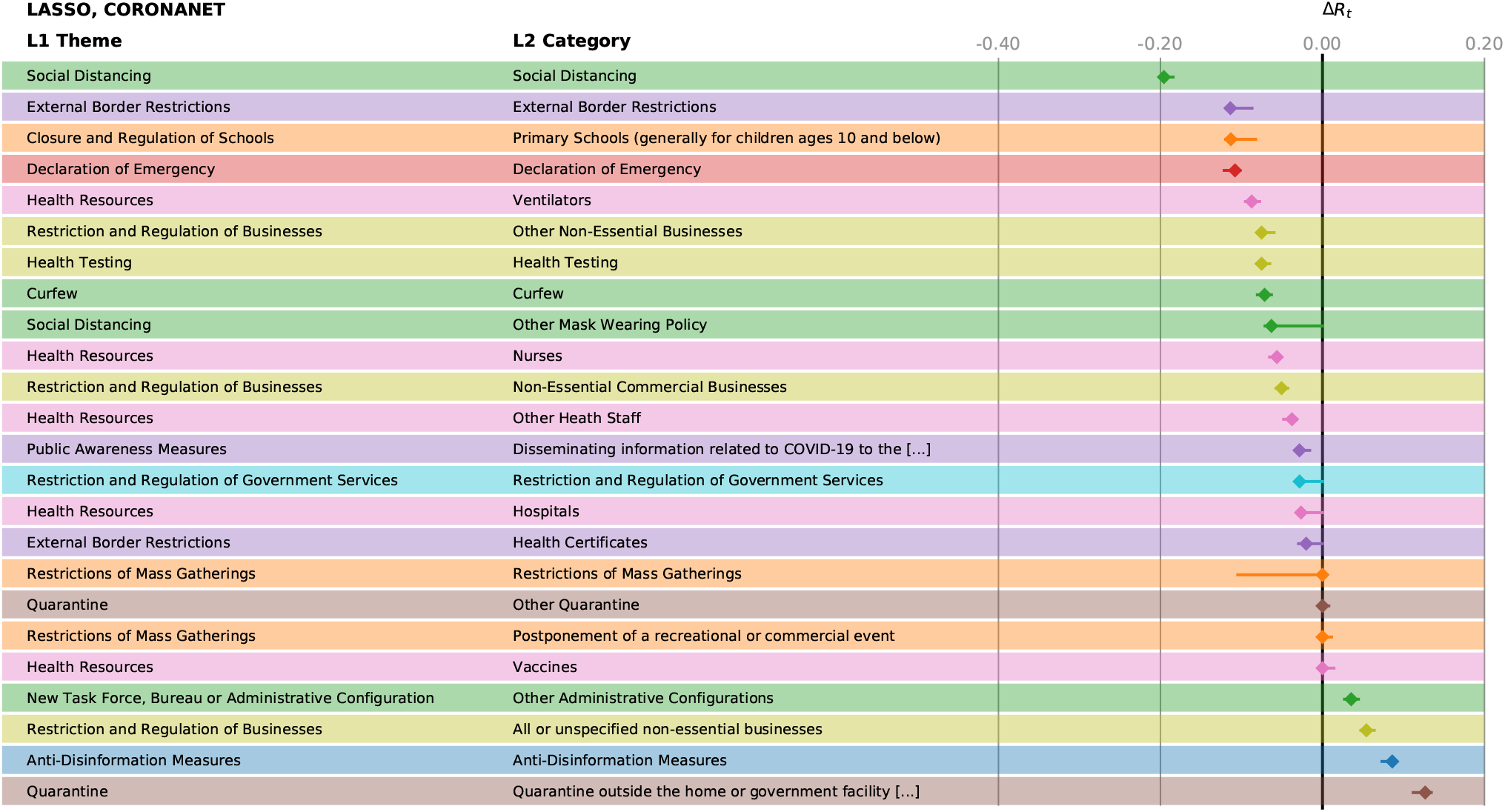
Effectiveness, Δ*R*_*t*_ of the different NPIs in the LASSO analysis for CORONANET. The horizontal bars mark the 95% confidence intervals.

**Figure S17:**
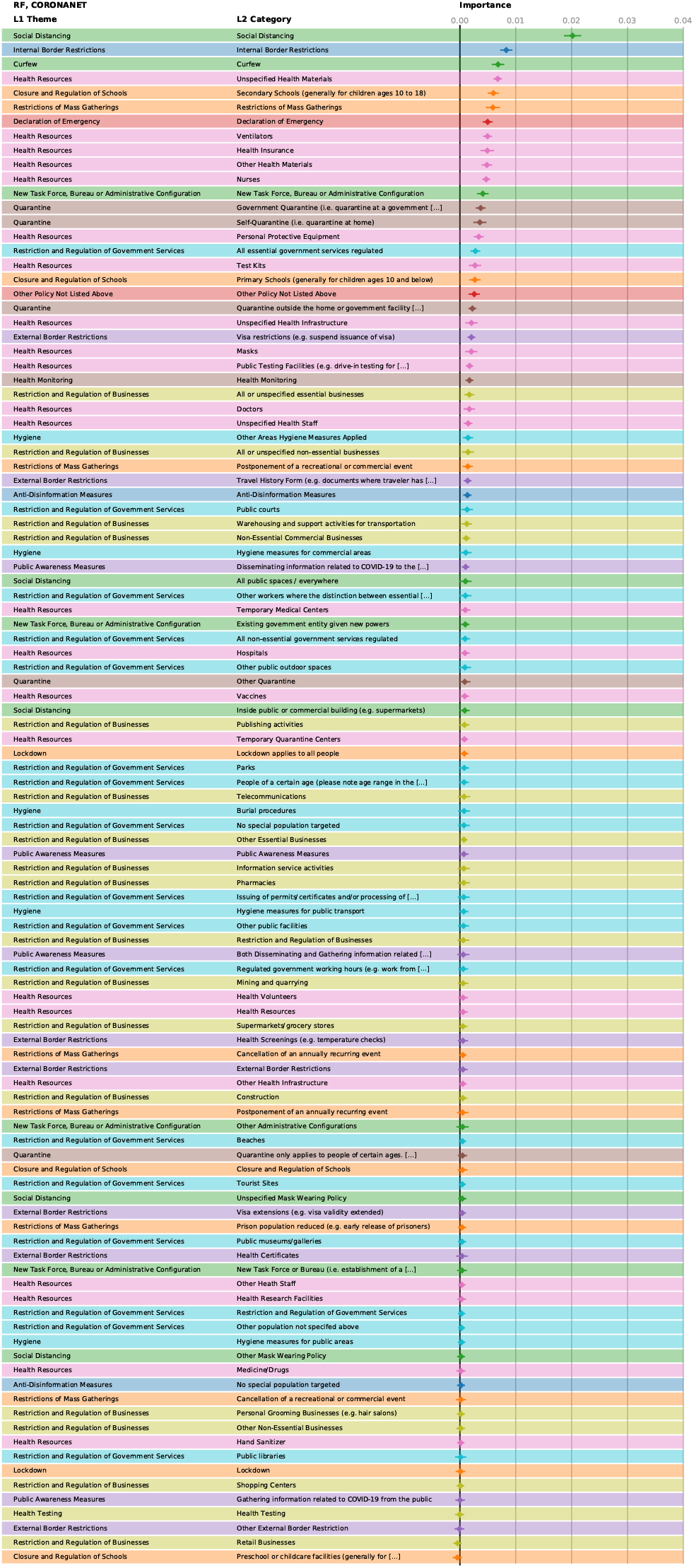
Feature importance of the different NPIs in the random forest model for CORONANET. The horizontal bars mark the 95% confidence intervals.

**Figure S18:**
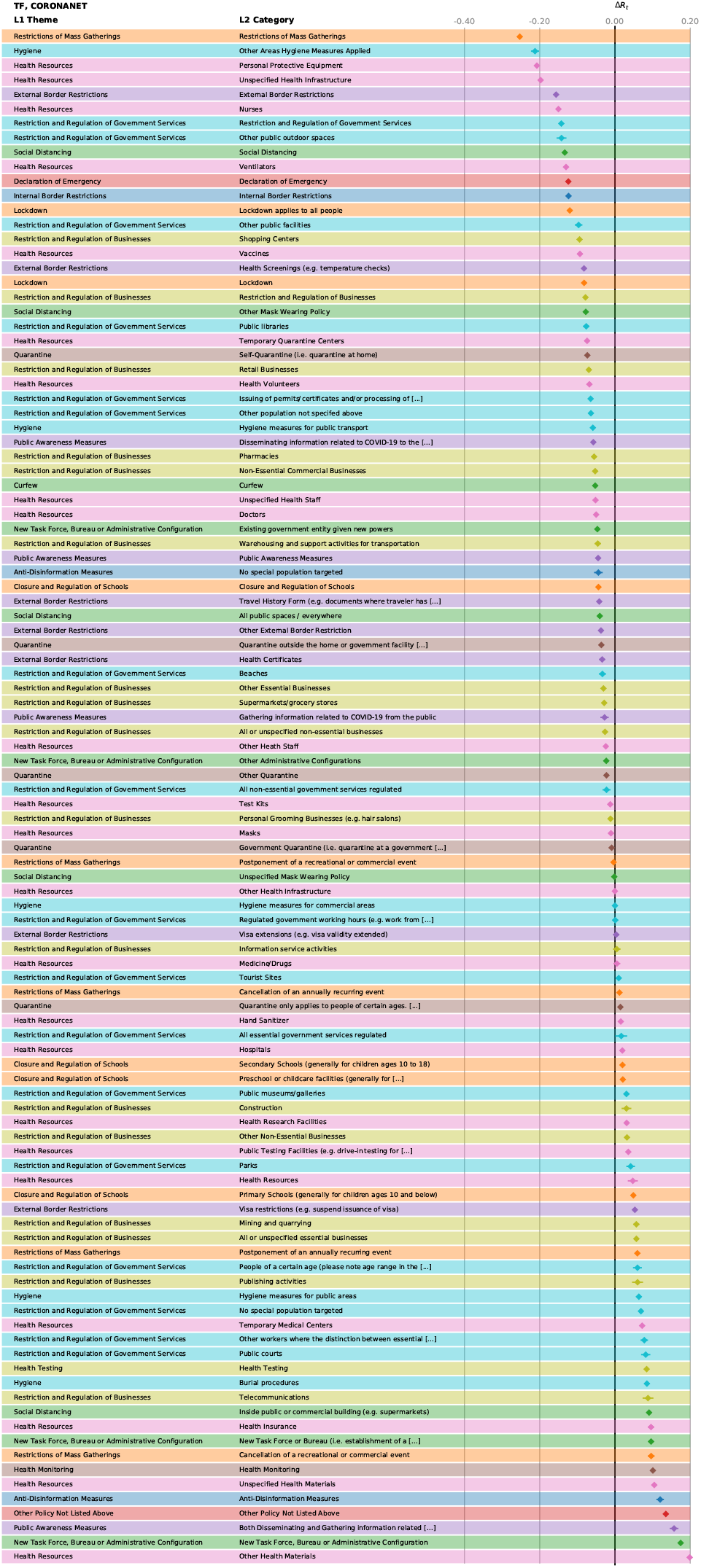
Effectiveness, Δ*R*_*t*_ of the different NPIs in the Transformer analysis for CORONANET. The bars marking the 95% confidence intervals are too small to be visible.

**Figure S19:**
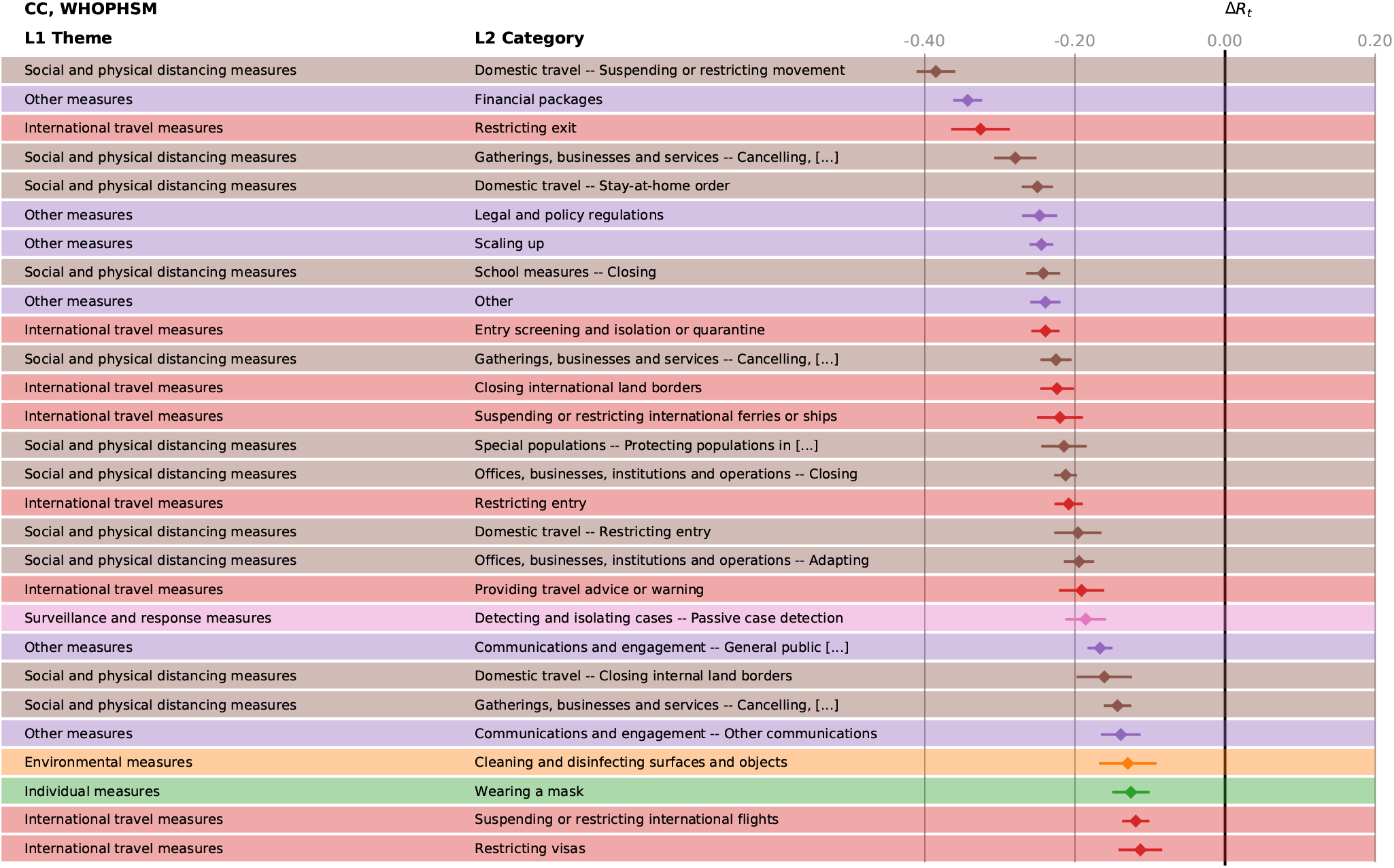
Effectiveness, Δ*R*_*t*_ of the different NPIs in the CC analysis for WHOPHSM. The horizontal bars mark the 95% confidence intervals.

**Figure S20:**
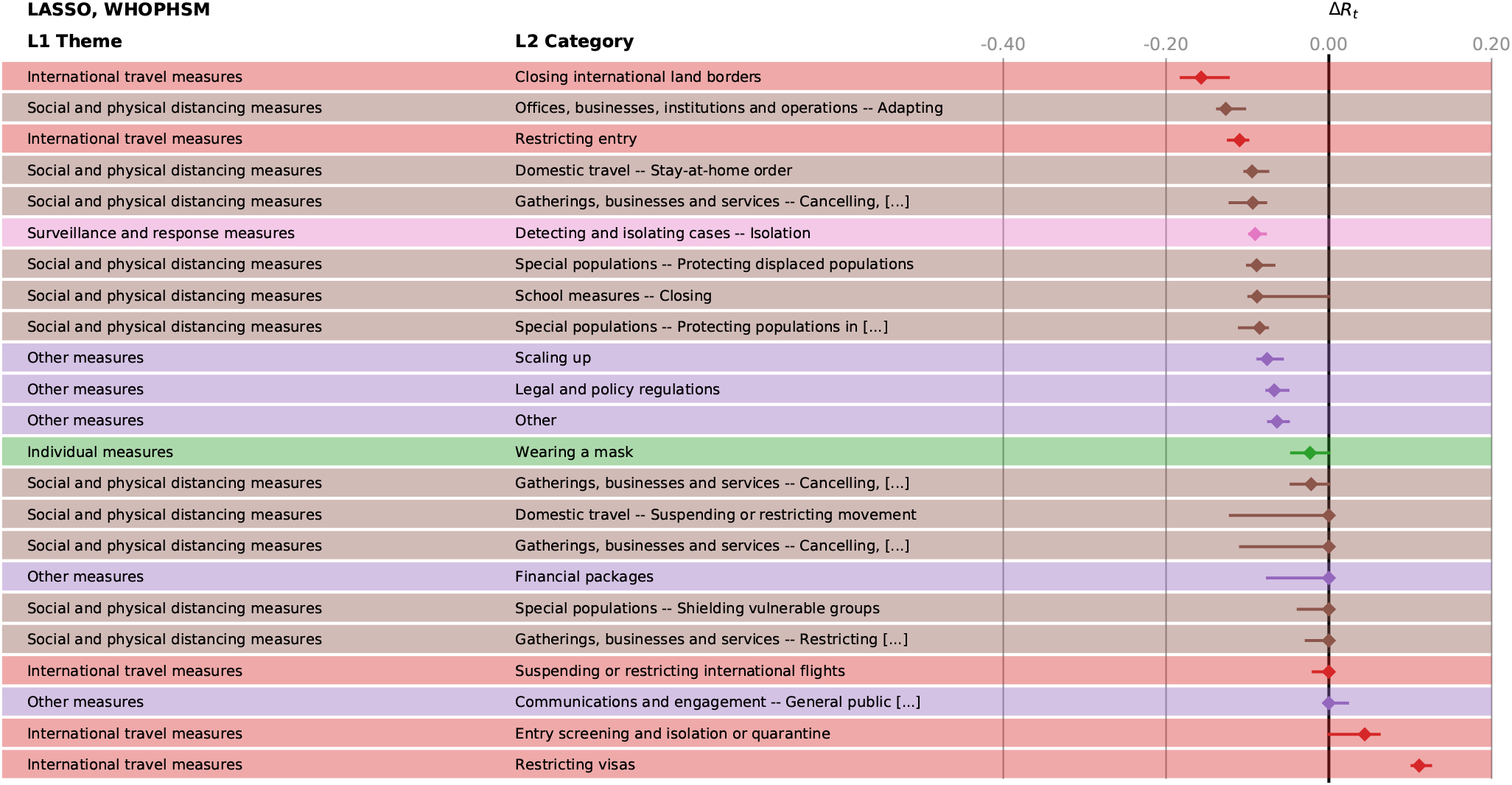
Effectiveness, Δ*R*_*t*_ of the different NPIs in the LASSO analysis for WHOPHSM. The horizontal bars mark the 95% confidence intervals.

**Figure S21:**
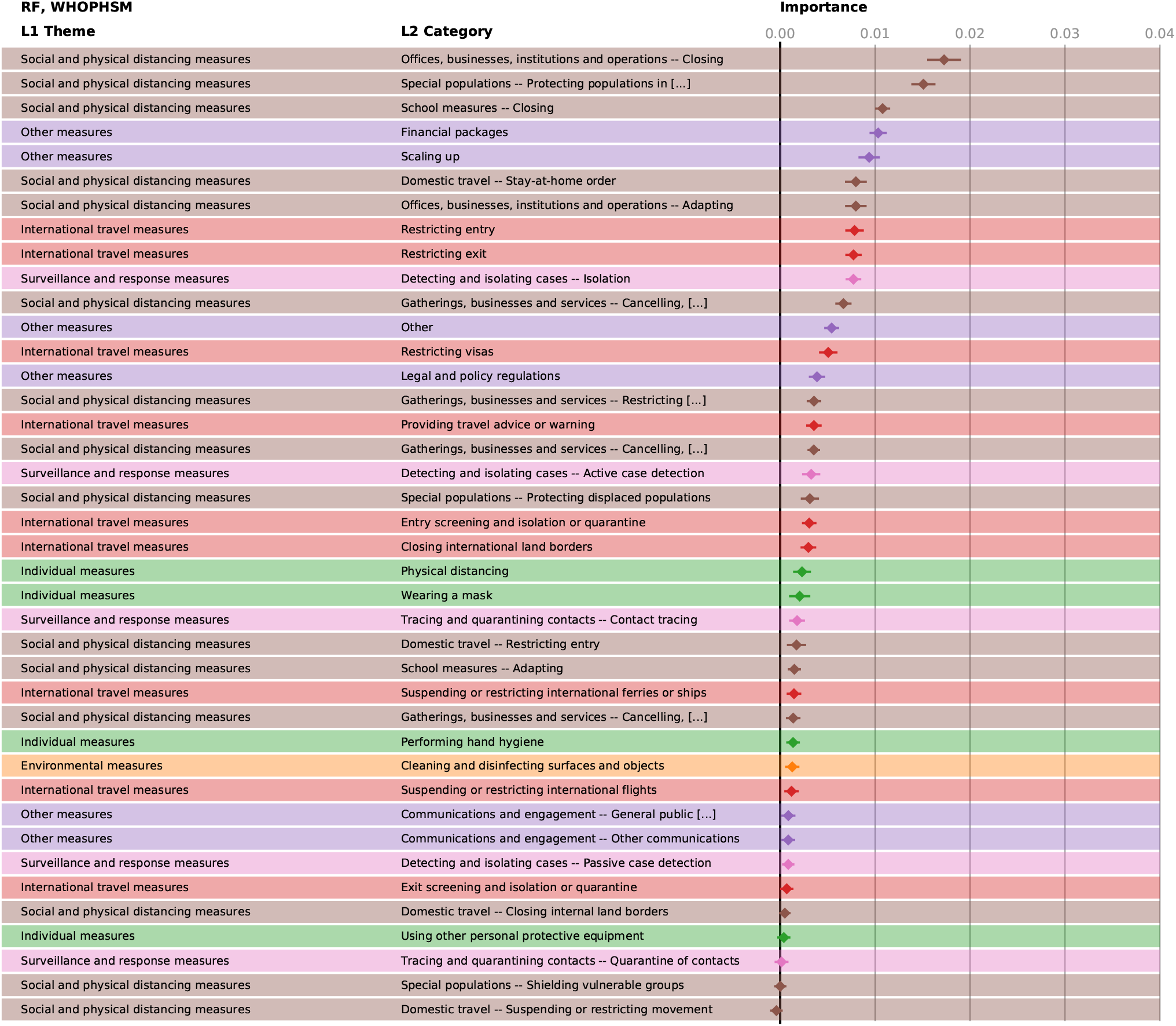
Feature importance of the different NPIs in the random forest model for WHOPHSM. The horizontal bars mark the 95% confidence intervals.

**Figure S22:**
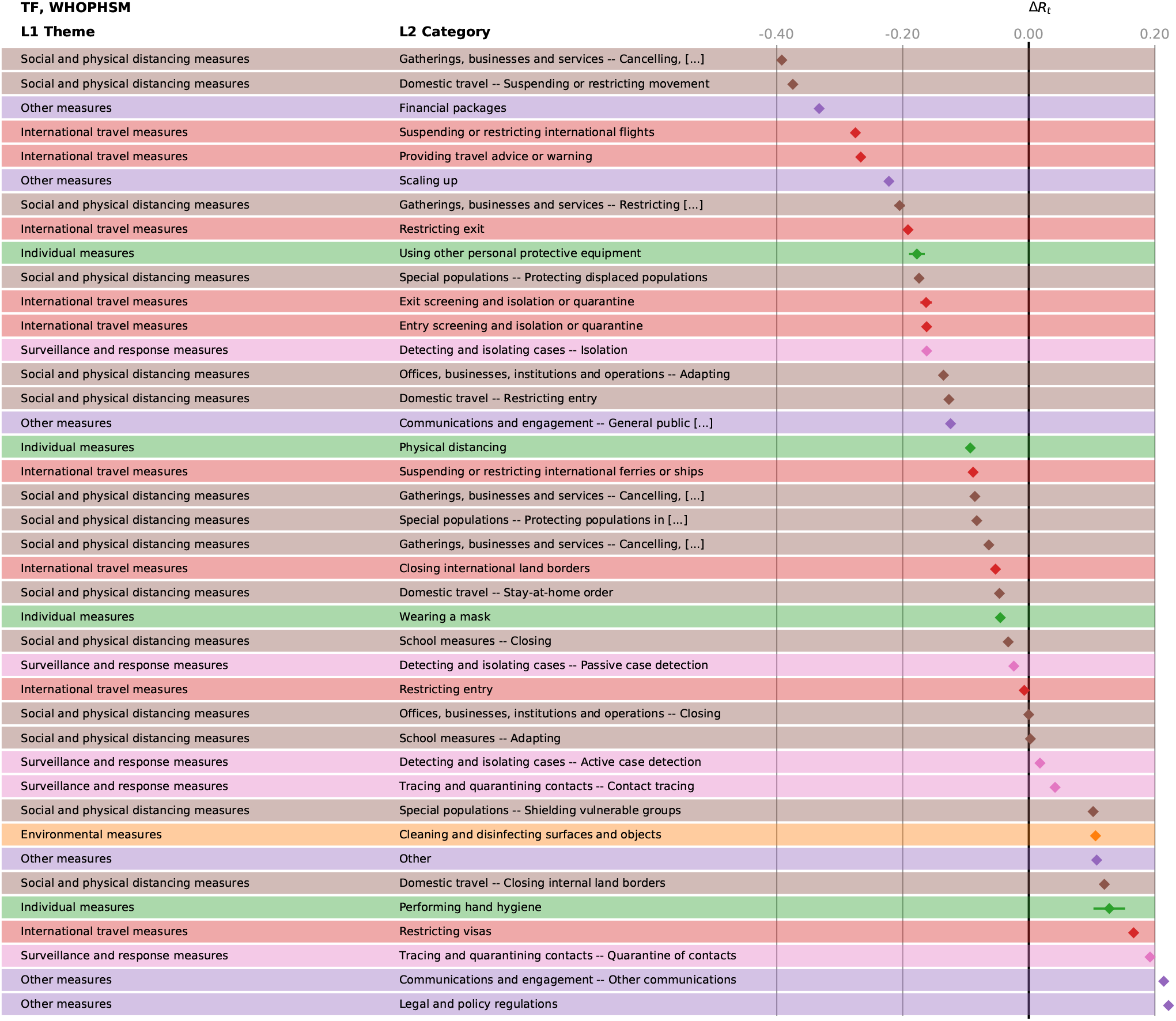
Effectiveness, Δ*R*_*t*_ of the different NPIs in the Transformer analysis for WHOPHSM. The bars marking the 95% confidence intervals are too small to be visible.

Two external datasets further corroborate our findings. These datasets also contain dedicated entries for policies related to mask wearing, which appear to show moderate reductions in *R*_*t*_ when compared to the other measures discussed above. We further confirmed the estimated impact of all consensus measures using two external datasets presenting a broader geographical coverage (and therefore counting more individual NPIs). This large analysis is, to date, unique, and proves the robustness of our results. It also shows that, although the NPI trackers have been built independently, sometimes for different purposes, and present different semantics and structure (which is a limitation to harmonizing results), their analysis provides convergent results with four methods, and the use of a smaller number of countries (56 in the CCCSL versus ∼ 200 in the CoronaNet and WHO-PHSM datasets) does not qualitatively affect the outcome. This finding is of importance when analysis of government policies needs to be conducted in emergency, to save computational time.

## 8 Discussion of results, organized by L1 theme

### Social distancing

Bans of small gatherings (gatherings of 50 persons or less) and the closure of educational institutions have a more substantial effect on *R*_*t*_ (but are also more intrusive to our daily lives) than the prohibition of mass gatherings, measures targeting special populations (e.g., elderly, vulnerable populations, hospitalised patients, prisoners or more exposed non-healthcare professionals) or adaptive measures for certain establishments (e.g., places of worship, administrative institutions, entertainment venues, nursing homes). While in earlier studies based on smaller numbers of countries, school closures had been attributed only a little effect on the spread of COVID-19 ^19, 20^, more recent evidence has been in favour of the effectiveness of this NPI ^28, 29^. This is also in line with a contact tracing study from South Korea which identified adolescents aged 10–19 as the biggest spreaders in household settings ^30^. Social distancing measures are less effective in countries with a high population density and a high degree of citizen participation in the government, as well as freedom of expression or free media (WGI Voice & Accountability). The country-level analysis confirms that these NPIs have a particularly high entropy, meaning that their effectiveness varies indeed substantially across countries. An exception to that are the measures for public transport and work safety protocols, where the latter mostly refers to mandatory guidelines for, e.g., physical barriers or fever checks at workplaces. These two social distancing measures have a low effectiveness rank (little significance across the methods) and low entropy, meaning that they had no impact on *R*_*t*_ consistently across most countries.

**Figure S23:**
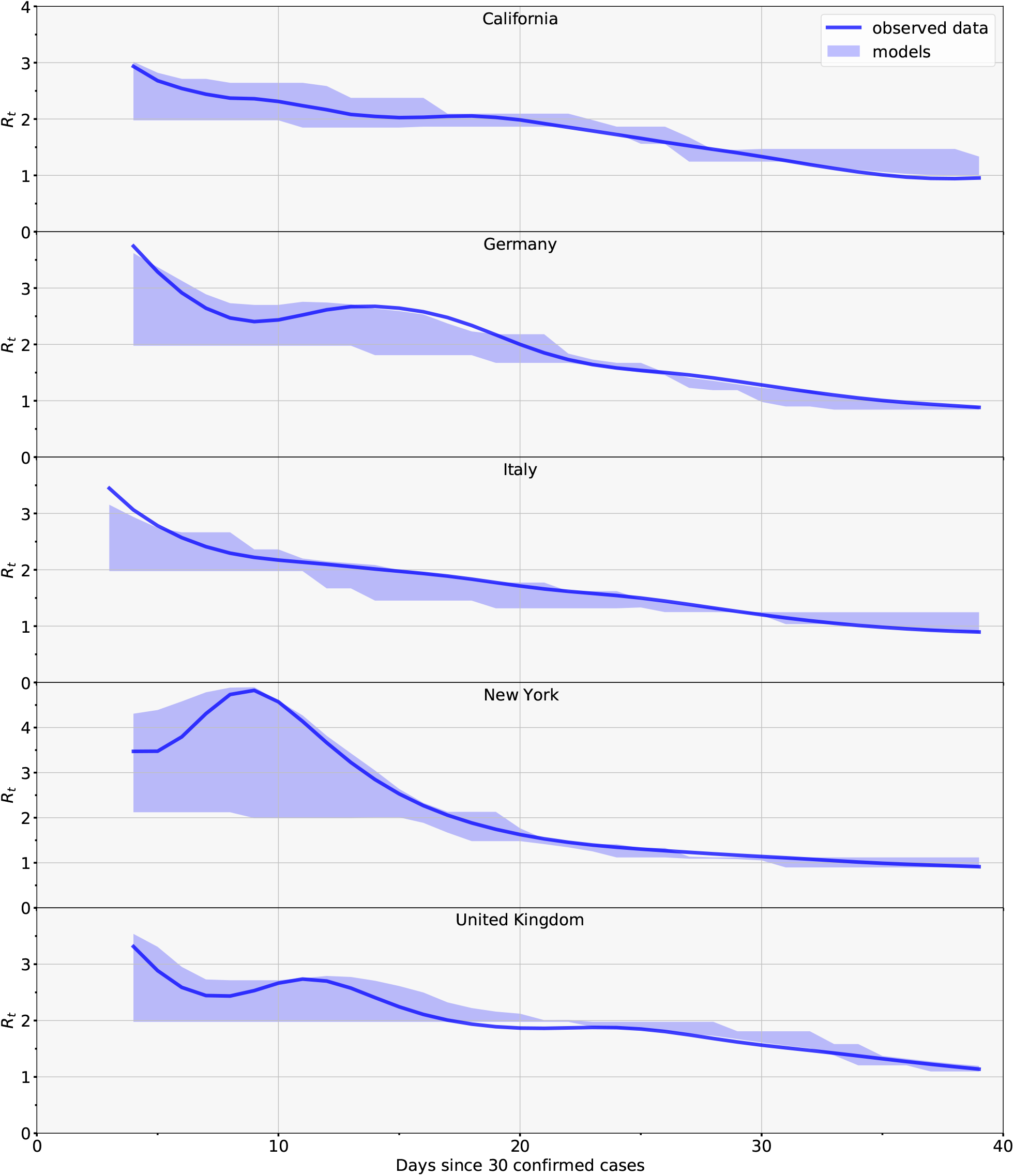
Observed values for *R*_*t*_ (solid lines), together with the range of values for *R*_*t*_ as predicted from the different methods (shaded regions).

**Figure S24:**
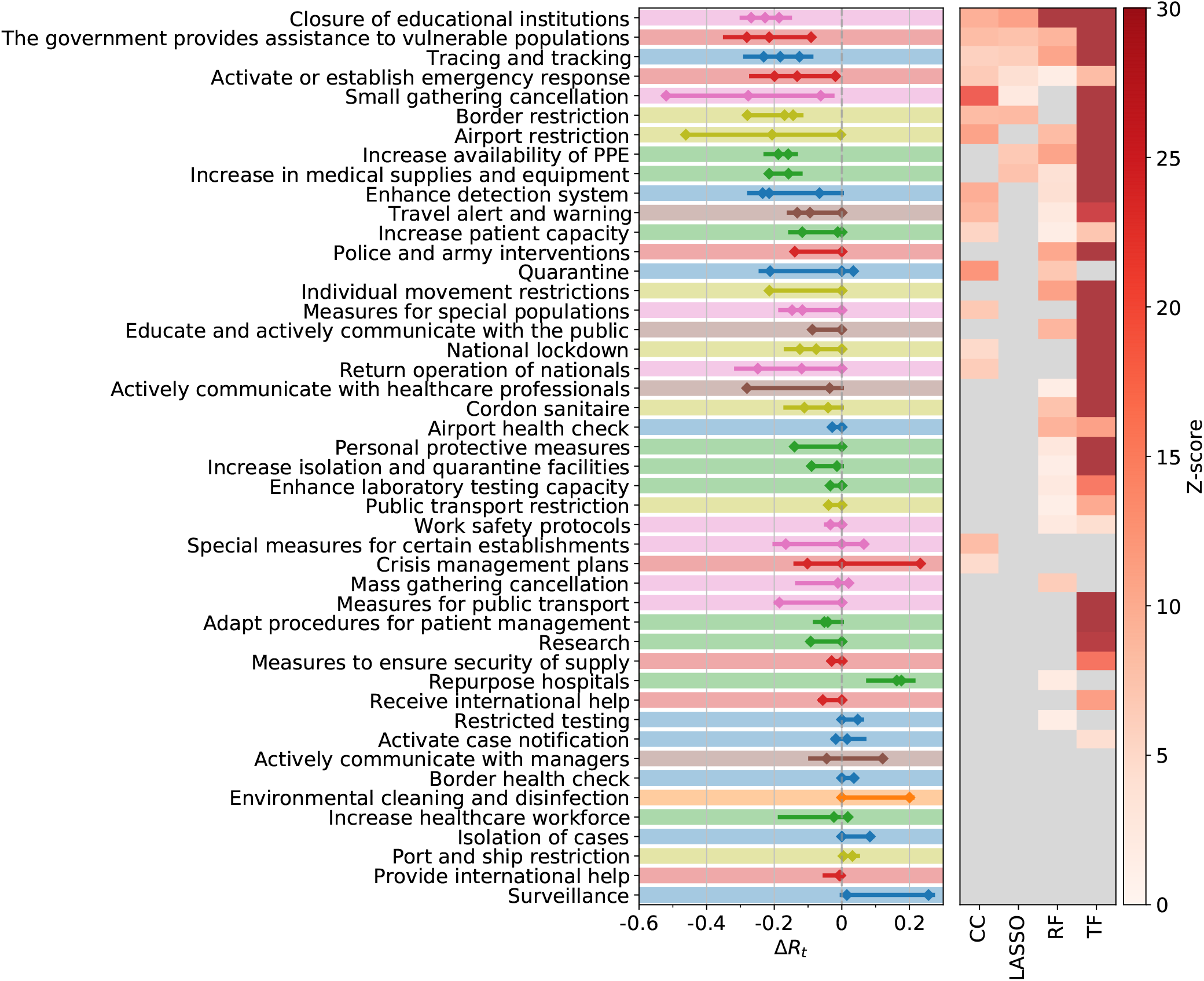
Analogue to Fig. 1 of the main text if the analysis is done on all countries except those in Europe and Africa.

**Figure S25:**
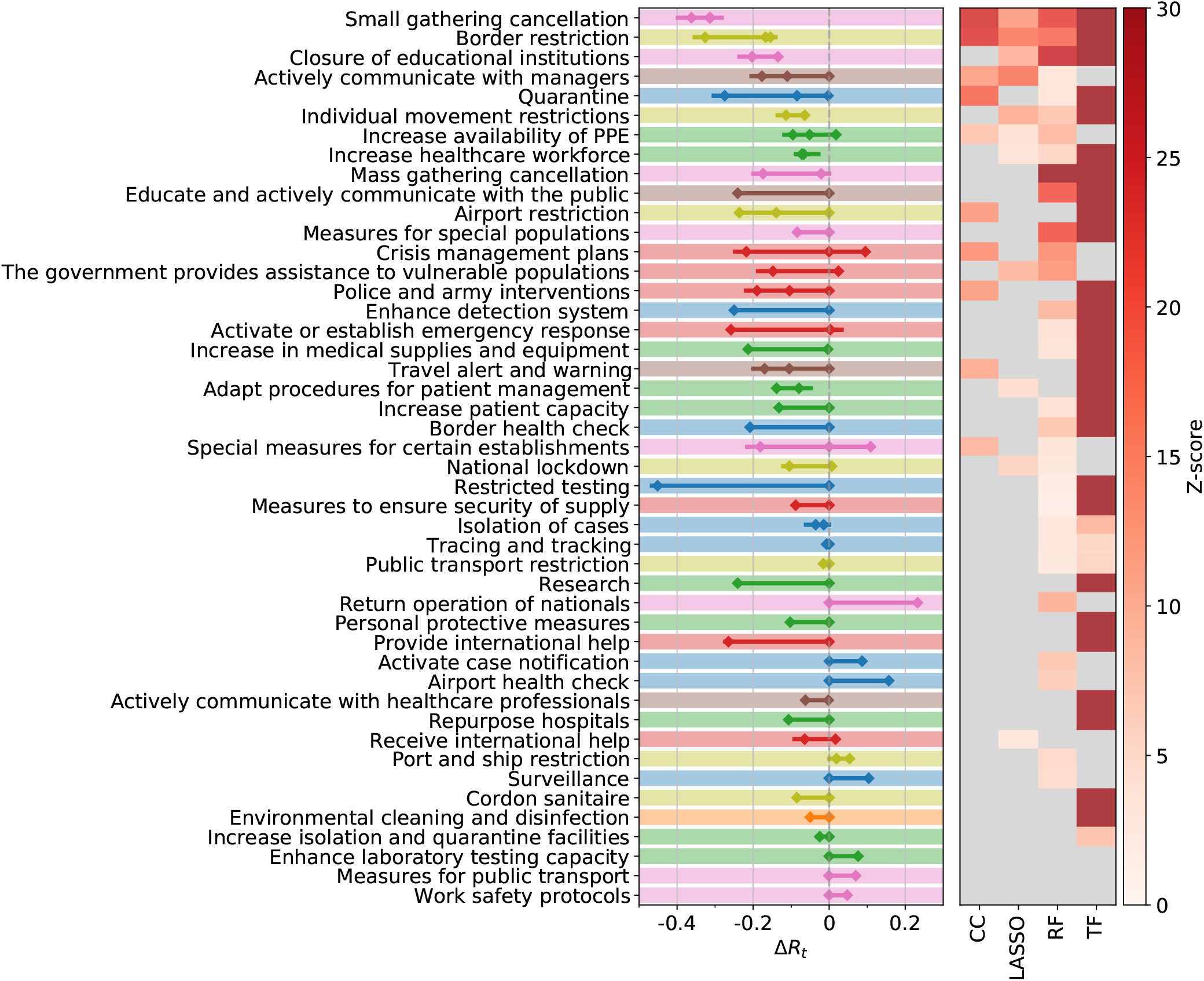
Analogue to Fig. 1 of the main text if the analysis is done on all countries except those in Asia and Oceania.

**Figure S26:**
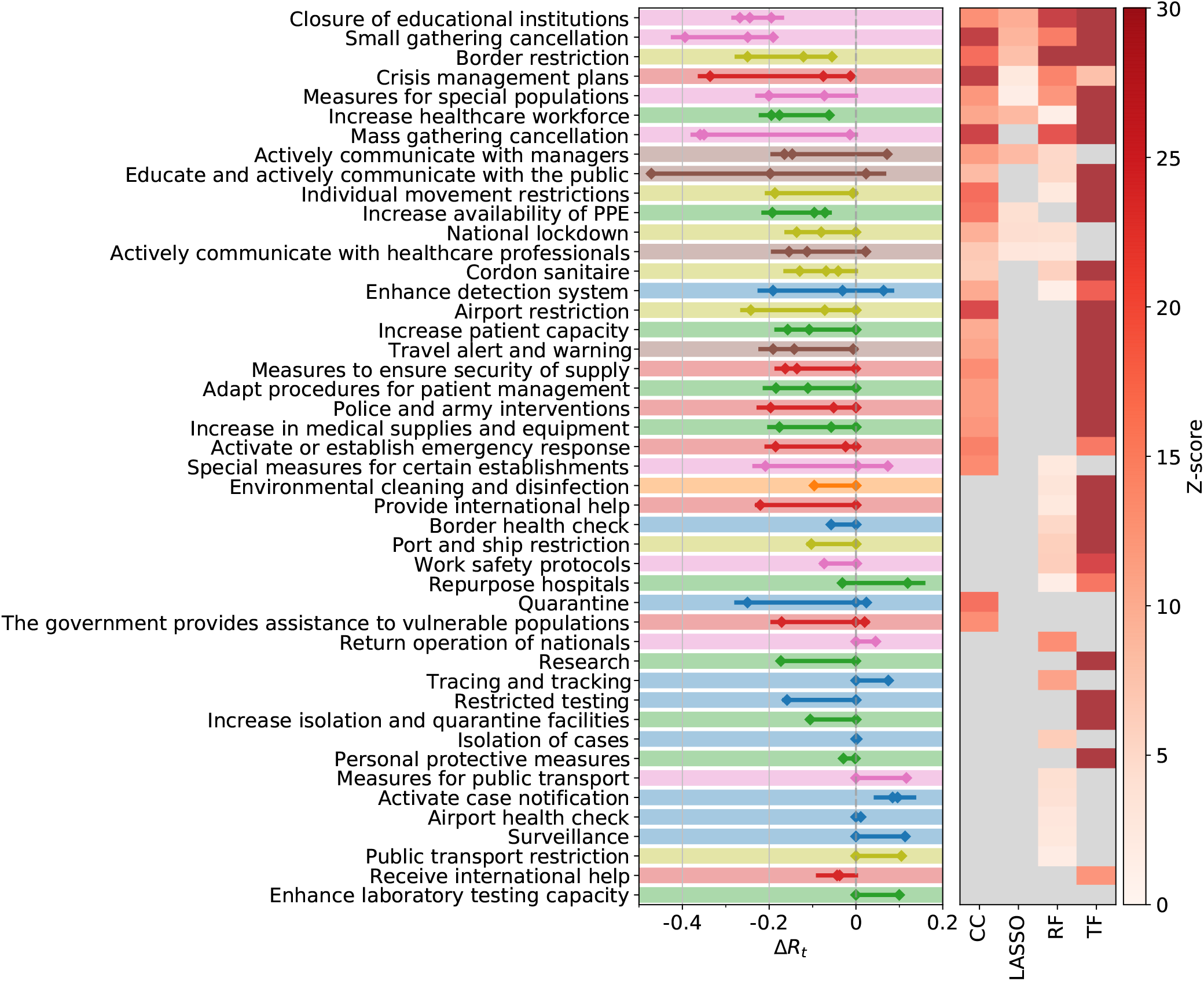
Analogue to Fig. 1 of the main text if the analysis is done on all countries except those in North, Central or South America.

**Figure S27:**
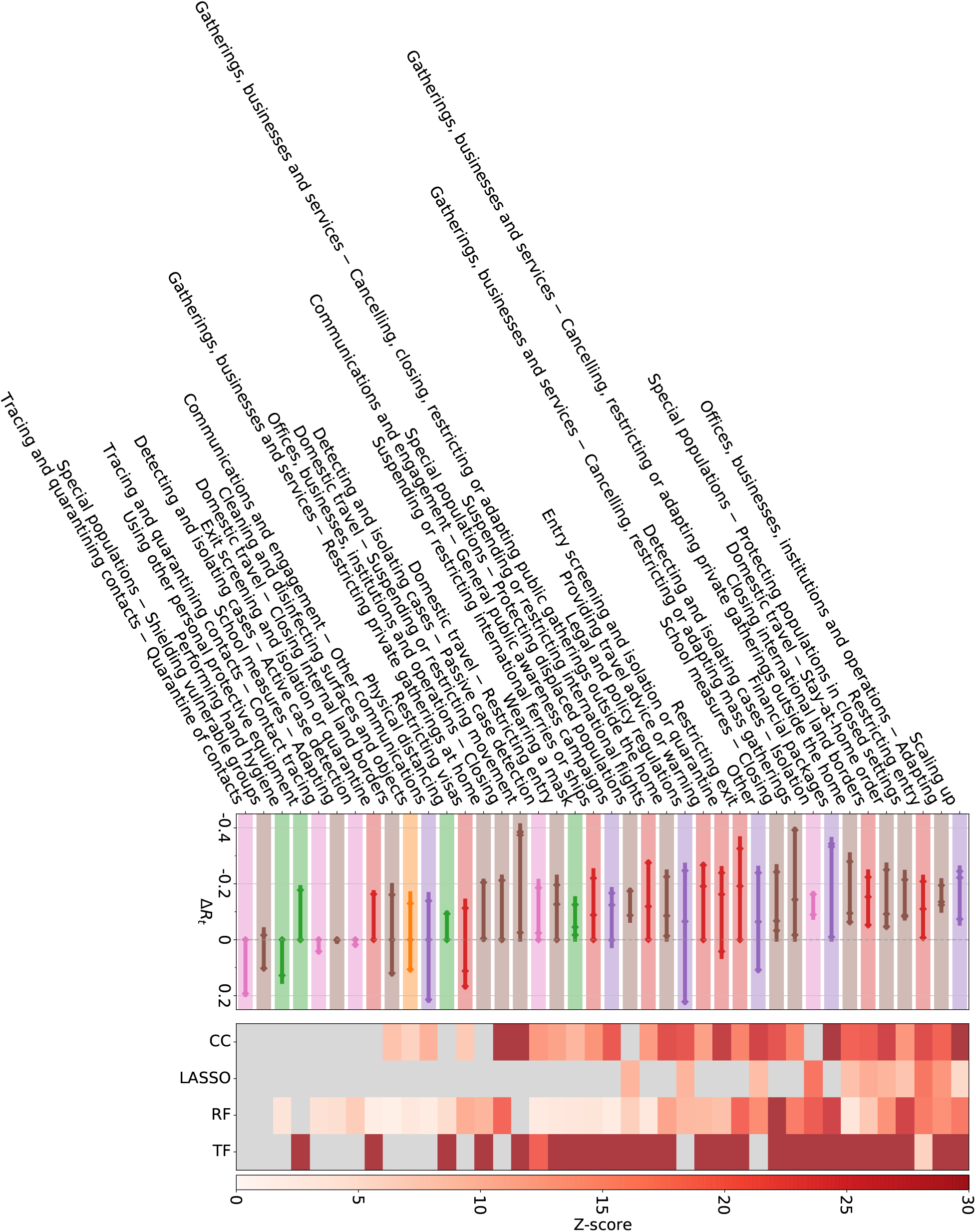
Analogue to Fig. 1 of the main text if the analysis is done on the WHOPHSM data set.

**Figure S28:**
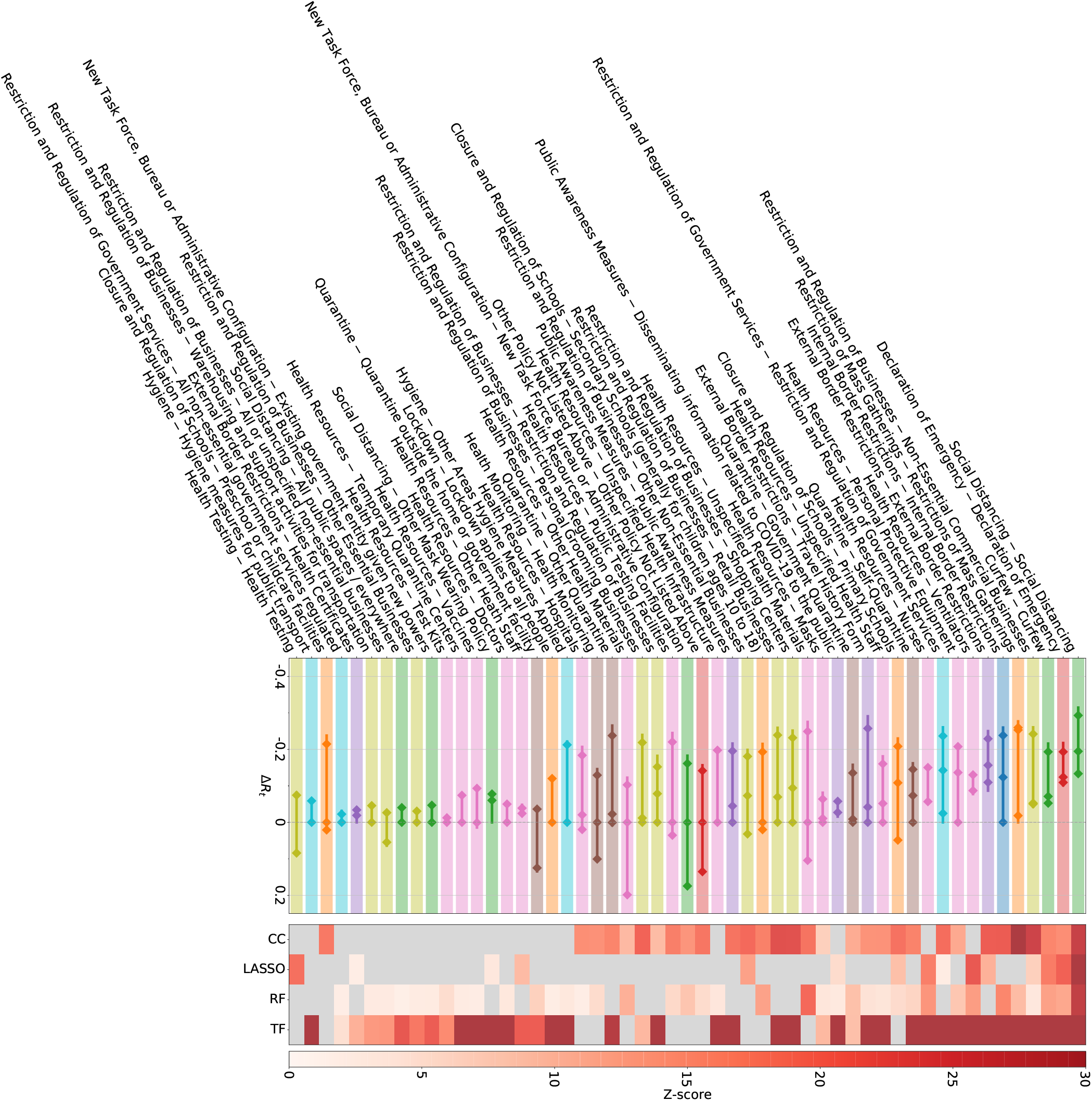
Analogue to Fig. 1 of the main text if the analysis is done on the CORONANET data set (continued on the next page).

**Figure S29:**
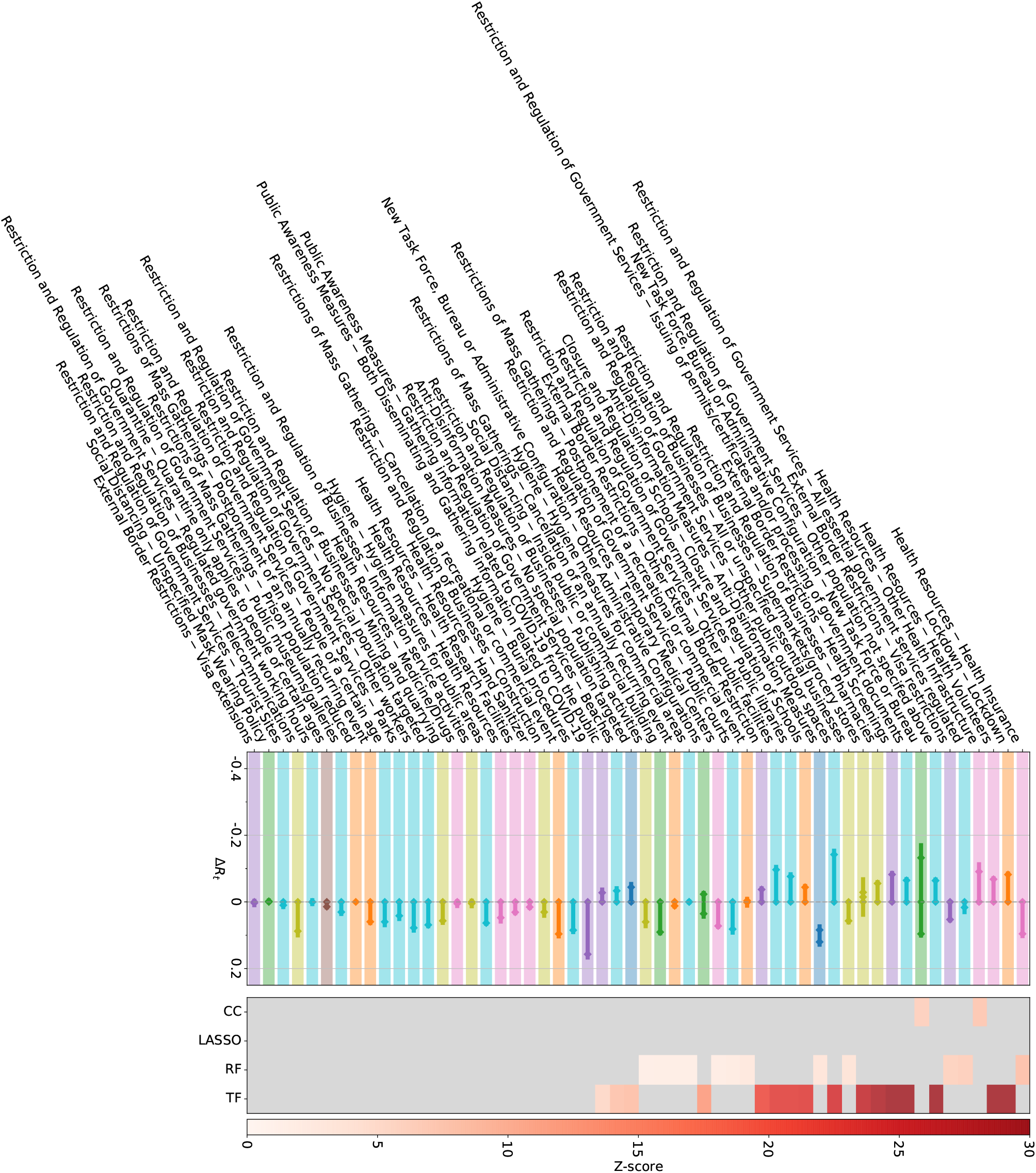
Analogue to Fig. 1 of the main text if the analysis is done on the CORONANET data set (continued).

### Healthcare and public health capacity

An increase in the availability of personal protective equipment (PPE) to the healthcare workforce, together with measures aiming to reduce the number of non-COVID-19 or non-critical COVID-19 patients in medical centres and hospitals (by promoting self-isolation of mildly symptomatic patients, setting up health hotlines, etc.) are also essential building blocks of successful containment strategies. All of these measures combine high effectiveness of early implementation and low entropy, meaning that they are similarly effective in most countries. Actions aiming to enhance the health system are critical. They are the primary response to patients and have no (or few) negative repercussions on individual rights of liberty (exception on the travel restriction for healthcare workers imposed by several countries). Our results demonstrate that government support to the health system needs to be a priority during a health crisis in order to reduce mortality ^66^. In line with our result “the earlier, the better”, we argue that those actions must be taken early enough to prepare for a surge in healthcare demand. Compared to other interventions, increased medical supplies and availability of PPE show substantially stronger positive correlations with several governance indicators including government effectiveness and control of corruption. Indeed, there are increased news reports currently on scandals related to government procurement of PPE ^67–69^.

### Travel restrictions

Different types of travel restrictions also show significant effects, in particular border restrictions (e.g., border closure, border controls), individual movement restrictions (e.g., curfews, the prohibition of non-essential activities) and cordons sanitaires (containment zones). The high effectiveness of border restrictions is driven by European countries (its impact on *R*_*t*_ turns insignificant in two of our methods after removing all European countries); most likely for geographic reasons. This finding is in line with a high entropy score of border, airport, port and ship as well as individual movement restrictions.

Effectiveness of ultimate measures such as stay-at-home orders or lockdowns is still controversial. Recent studies suggest that a national lockdown reduces *R*_*t*_ by an average of 5% ^19^ to 80% ^20^, whereas other interventions seem to reduce the virus spread by 5% ^20^ to 30% ^19^. In some countries or territories, the effect of a lockdown decided in the late stage of the epidemic may not be more effective than previously implemented bans on gatherings ^19, 20, 70^. Our analysis highlights the importance of early national lockdowns by showing how the relative effectiveness of that measure correlates with the epidemic age of its adoption. However, the reduced effectiveness of lockdowns at higher epidemic age, as observed in Fig. 4, does not necessarily imply that taking this NPI late is useless.

### Risk communication

In terms of risk communication, we find that pro-active communication with stakeholders from the private sector (e.g., business owners or chief executive officers) to promote voluntary safety protocols in enterprises, businesses, event organization, government administrations, etc., shows a significant effect in each of the four analyses, mainly when implemented early. Three out of four approaches also indicate a substantial impact of public health communication strategies (i.e., non-binding NPIs) encouraging citizen engagement and empowering them with information.

### Resource allocation

Measures for resource allocation show limited impacts on *R*_*t*_ in our analysis (e.g., police and army interventions being insignificant in all studies) with relatively high entropy, meaning that country-level effects are important. Surprisingly, the implementation of crisis management plans turns out to be highly effective, except for the Americas. After removing countries from North and South America from the analyses, all four of our methods agree on significant effects of crisis management plans with an Δ*R*_*t*_ of down to −0.3, suggesting a lack of *effective* crisis plans in American territories. For instance, US states had to focus on providing health insurance and economic stimulus as well as facilitating administrative procedures, while European countries could develop their plans on top of a stronger socio-economic basis ^71, 72^. Crisis management plans are also more effective in countries with a non-participatory government, meaning that countries with increasingly authoritarian practices might be at an advantage at implementing such policies, as can be seen in the swift response of Singapore ^73^.

### Case identification, contact tracing and related measures

NPIs related to case identification and contact tracing show some of the lowest effectiveness ranks and in some cases even increase *R*_*t*_, consistently across most countries (NPIs with the five lowest entropy scores all belong to this theme). This result is to be expected, as, e.g., increased testing and faster contact tracing will on the short-run increase the numbers of found cases in return for reduced numbers in the long run. We do not assess such long-term effects (over timespans of more than a month) in the current work. Furthermore, note that our analysis considers mostly data from March and April 2020 where many countries experienced surges of case numbers that most likely hindered effective contact tracing and other case identification measures. This also applies to the relative ineffectiveness of quarantining people who either are infected or were exposed to infected persons, while the promotion of self-isolation of people with symptoms was one of the most effective NPIs. This result confirms a tendency in our results where voluntary measures are more effective than similar mandatory ones.

## 9 Additional tables

**Table S3:** Subcategories (L3) belonging to the eight consensus categories (L2) identified as significant by all four methods.

| L2 category | L3 subcategories |
| --- | --- |
| Small gathering cancellation | Complete prohibition of gathering; Closure of restaurants/bars/cafes; Closure of non-essential shops; Limit up to 5 persons; Implement part-time work; Mandatory home office; Limit up to 10 persons; Restriction on private and familial events; Limit up to 30 persons; Limit up to 2 persons; Remote Psychotherapy Consultation; Limit up to 6 persons; Limit up to 20 persons; Closure of short-term accommodation; Limit up to 1 persons; Closure of student dormitories; Reduce close physical contact in workplaces; Mandatory 2m distance in public spaces; Limit up to 3 persons; Limit up to 50 persons; Limit up to 4 persons; Limit up to 25 persons; Non-critical court operations suspended |
| Closure of educational institutions | Complete closure of kindergartens; Complete closure of primary and secondary schools; Complete closure of universities; Reduction of excursions, out-of-house events; Complete closure of all educational institutions; Cancellation of exams; Partial closure of primary and secondary schools; Partial closure of universities; Complete closure of secondary schools; Extracurriculars cancelled; Partial closure of educational institutions; Rules for exams; Partial closure of kindergartens; Restrictions on exams; Closure of adult educational schools; Complete closure of scientific institutions; Complete closure of research institutes |
| Border restriction | Land borders closed; Land borders closed; Entry ban to people from high-risk areas other than China; Land border controls; Entry ban to non-citizens; Conditional entry of persons from neighboring countries; Conditional entry of citizens; Entry ban to non EU citizens; Temporary reduction of service; Entry ban to people from China; Entry ban to refugees; Entry ban for symptomatic people and case contacts; Travel ban to high-risk areas; Total entry ban; Close land border to prevent virus spread; Entry ban to infected persons; Entry ban to people with a travel history to China; Ban on passenger transport from China; Force departure of Chinese nationals; Border control; Suspension of passenger railway transports crossing the country border; Restriction of freight transport; Ban on road passenger transport from high risk areas |
| Increase availability of PPE | PPE for healthcare professionals; Face masks; PPE (not specified); PPE other than face masks; Prohibition of export of protective personal equipment; Hand sanitizers; Increase domestic production of PPE |
| Individual movement restrictions | Movements for non-essential activities forbidden; Curfew; Non-essential travels abroad/out-of-state forbidden; Segmentation of the population; Partial restriction on movements; Prohibition of moving out the municipality of residence; Restrictions on the movements of children |
| National lockdown | for 21 days; For 2 weeks; Stay-at-home Order; Safer-at-home Order |

**Table S1:** Number of consensus NPIs after removal of the indicated continents, compared to the expected number of consensus NPIs if the significance of the NPIs in the different methods was statistically independent.

| Continents | Number of consensus measures | Expectation value |
| --- | --- | --- |
| Europe and Africa | 4 | 1.05 |
| Asia and Oceania | 2 | 1.05 |
| Americas | 6 | 2.40 |

**Table S2:** Results of the CC analysis for risk communication NPIs on level L3. For each measure we give the change in *R*_*t*_ with the SE in brackets and its *p*-value. Measures with significant effects after a multiple testing correction are highlighted in bold.

| L3 category | L2 category | $\Delta R_t$ (SE) | $p$-value |
| --- | --- | --- | --- |
| **Warning against travel to and return from high risk areas** | **Travel alert and warning** | **-0.14(1)** | **< 10<sup>-7</sup>** |
| **Encourage stay at home** | **Educate and actively communicate with the public** | **-0.14(1)** | **< 10<sup>-4</sup>** |
| **Promote social distancing measures** | **Educate and actively communicate with the public** | **-0.20(2)** | **< 10<sup>-4</sup>** |
| **Promote workplace safety measures** | **Educate and actively communicate with the public** | **-0.19(2)** | **< 10<sup>-4</sup>** |
| **Promote self-initiated isolation of people with mild respiratory symptoms** | **Educate and actively communicate with the public** | **-0.19(2)** | **0.0001** |
| **Information campaign** | **Educate and actively communicate with the public** | **-0.13(2)** | **0.0003** |
| Respiratory etiquette | Educate and actively communicate with the public | -0.10(1) | 0.0005 |
| Answer to questions | Educate and actively communicate with the public | -0.077(1) | 0.0007 |
| Call for return of nationals living abroad | Educate and actively communicate with the public | -0.25(2) | 0.0009 |
| Recommendations for work safety protocols | Actively communicate with managers | -0.13(2) | 0.0013 |
| Encourage self-initiated quarantine | Educate and actively communicate with the public | -0.19(2) | 0.005 |
| Communication targets protection of vulnerable populations | Educate and actively communicate with the public | -0.14(2) | 0.007 |
| Guidelines | Actively communicate with managers | -0.14(2) | 0.02 |
| Information about travels | Educate and actively communicate with the public | -0.17(3) | 0.02 |
| Direct government communication | Educate and actively communicate with the public | -0.072(2) | 0.02 |
| Encourage hand hygiene | Educate and actively communicate with the public | -0.084(2) | 0.03 |
| Direct advice to vulnerable populations | Educate and actively communicate with the public | -0.14(3) | 0.04 |
| Foster community assistance | Educate and actively communicate with the public | -0.14(2) | 0.04 |

